# Genetic Prediction of Circulating Lipoprotein(a) Levels in Diverse Populations

**DOI:** 10.64898/2026.02.20.26346738

**Authors:** Michael G. Levin, Margaret Sunitha Selvaraj, Ha My T. Vy, Renae Judy, Michael C. Honigberg, Archna Bajaj, Girish Nadkarni, Ron Do, Pradeep Natarajan, Joshua C. Denny, Po-Ru Loh, Penn Medicine Biobank

## Abstract

**Background:** Circulating lipoprotein(a) [Lp(a)] levels are highly heritable and linked to atherosclerotic cardiovascular disease, yet clinical measurement rates remain low (<1%) in the United States. The high heritability of Lp(a) across populations makes genetic prediction an attractive approach for closing this testing gap, but existing polygenic scores transfer poorly across populations. Haplotype-based prediction models, which use standard genome-wide genotype data to capture common-, rare-, and structural-variation at the LPA locus, could bridge this gap, enabling opportunistic identification of individuals with elevated Lp(a) levels across diverse populations within existing large, genotyped cohorts.

**Objectives:** This study sought to develop and validate a haplotype-based prediction model using genome-wide genotype data to identify individuals with elevated Lp(a) levels across diverse populations.

**Methods:** We developed an *LPA*-haplotype model using data from the All of Us Research Program and validated it in the Penn Medicine BioBank (PMBB), Mass General Brigham Biobank (MGBB), and Mount Sinai BioMe cohorts. Primary outcomes included model performance for predicting continuous Lp(a) concentrations (r²) and identifying elevated Lp(a) levels (>125 nmol/L) through positive predictive value (PPV) and number needed to test (NNT).

**Results:** Among PMBB (n = 1856), MGBB (n = 1401), and BioMe (n = 1686) participants with available genotype and Lp(a) measurements, average age was 60 years, and 51% were female. Overall r² of the haplotype model was 0.46 (95% Credible Interval [CrI] 0.32 to 0.6), with similar performance across genetically inferred ancestries and cohorts. For identifying elevated Lp(a) levels >125 nmol/L the overall PPV was 0.81 (95% CrI 0.6 to 0.89), corresponding to a NNT of 1.2 (95% CrI 1.1 to 1.7) individuals predicted to have elevated levels needing to undergo clinical testing to identify one true elevation. In the full PMBB cohort (n = 49310), the haplotype model identified elevated Lp(a) at a rate of 128 per 1000 (95% CrI 125 to 130), corresponding to an estimated 14.4-fold improvement (95% CrI 13.1 to 15.9; P(improvement) = 1) in identification rate compared with the existing rate of clinical assessment.

**Conclusions:** A haplotype-based genetic model effectively identified individuals with elevated Lp(a) levels across diverse populations, with potential utility for opportunistic screening among cohorts where genotype data is available, but Lp(a) testing rates are low.

## INTRODUCTION

Circulating levels of lipoprotein(a) Lp(a) are highly heritable, with 70–90% of interindividual variation attributable to genetic variation at the LPA locus on chromosome 6.^1^ Despite robust links to atherosclerotic cardiovascular disease and guideline recommendations for one-time testing in adults, rates of clinical Lp(a) measurement remain below 1% in the United States.^2,3^ This testing gap has significant implications as Lp(a)-lowering therapies approach clinical availability, with multiple agents in late-stage development including antisense oligonucleotides and small interfering RNAs.

Given the high heritability of Lp(a) across populations and increasingly widespread genotyping, genetic prediction has been proposed as a strategy to identify individuals with elevated levels (>125 nmol/L) who have not already been assessed clinically.^4–6^ However, traditional polygenic scores (PGS) for Lp(a) have shown relatively reduced performance in non-European populations despite some of these groups—particularly individuals of African ancestry—having substantially higher rates of clinical Lp(a) elevations.^7,8^ This limitation reflects a fundamental challenge: PGS assume that the same genetic variants have consistent additive effects across populations, an assumption that breaks down for complex loci like LPA.

Recently, a highly-accurate Lp(a) prediction model was developed using whole-genome sequencing data from the UK Biobank.^1^ This sequencing-based model captures the kringle IV type 2 (KIV-2) repeat polymorphism, a primary determinant of circulating Lp(a) levels, as well as common and rare variation, importantly demonstrating consistent performance across diverse populations. However, clinical implementation is limited by the cost of sequencing (hundreds of dollars per sample compared with tens of dollars for genotyping) and the need for specialized computational pipelines.

To address this limitation, we derived a haplotype-based prediction model that utilizes inexpensive genome-wide genotype data already available for tens of millions of individuals through research biobanks and direct-to-consumer genetic testing. Rather than summing effects of variants across the genome as in traditional PGS, our approach identifies individuals who share specific chromosomal segments flanking the LPA locus with reference individuals whose Lp(a) levels are known. This approach captures the combined effects of common variants, rare variants, and structural polymorphisms without requiring whole-genome sequencing, enabling accurate but inexpensive prediction of Lp(a) levels.^9^ Leveraging diverse cohorts like All of Us to generate the haplotype reference panel may overcome traditionally limited cross-population performance of existing Lp(a) polygenic scores. Unlike broad population-based laboratory Lp(a) measurement, which requires a blood draw and specialized assay, haplotype-based prediction can be applied retrospectively to millions of individuals with existing genotype data, prioritizing those who would benefit most from confirmatory testing and clinical follow-up.

We applied this haplotype-based model to the Penn Medicine Biobank (PMBB), Mass General Brigham Biobank (MGBB), and Mount Sinai BioMe cohorts, representing three diverse populations with clinical and genetic data. We aimed to: 1) assess the performance of the haplotype model in predicting circulating Lp(a) levels and identifying individuals with elevated Lp(a), 2) estimate the number of individuals needing clinical testing to identify one true elevation, and 3) compare opportunistic genetic screening to existing clinical practice for identifying elevated Lp(a) in biobank settings.

## METHODS

An overview of the study is presented in **eFigure 1**.

**Figure 1:**
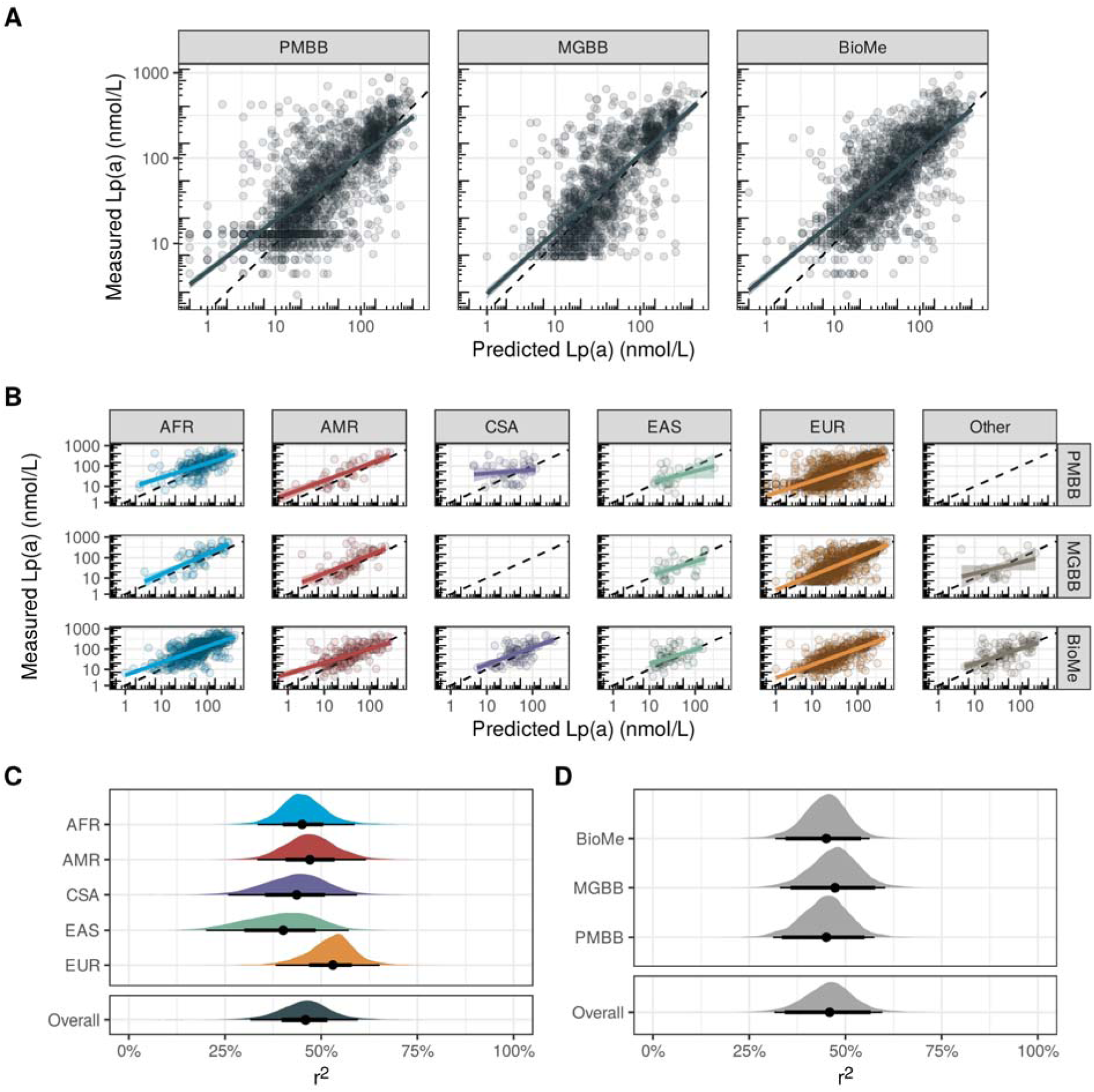
Measured vs. Predicted Lp(a) Concentrations in PMBB, MGBB, and BioMe. A) Scatterplots comparing clinically measured Lp(a) concentrations (y-axis) with haplotype-predicted concentrations (x-axis) across three validation cohorts: Penn Medicine BioBank (PMBB), Mass General Brigham Biobank (MGBB), and Mount Sinai BioMe. Dashed diagonal lines indicate perfect concordance. Both axes displayed on logDD scale. B) Performance stratified by genetically inferred ancestry and cohort. C) R² (variance explained) of the haplotype model stratified by genetically inferred ancestry, combining data across all three validation cohorts. D) R² stratified by cohort. Distributions represent posterior estimates from Bayesian mixed models with random effects for ancestry and cohort; points and horizontal lines indicate posterior medians and 95% credible intervals. AFR = African; AMR = Admixed American; CSA = Central/South Asian; EAS = East Asian; EUR = European.

### Ethical Approval

The Penn Medicine Biobank is approved by the University of Pennsylvania Institutional Review Board (IRB protocol #813913) and all participants provided written informed consent. The All of Us research program, Mass General Brigham Biobank, and Mount Sinai BioMe biobanks are approved by their respective institutional review boards and all participants provided written informed consent.

### Haplotype-Model Development

A haplotype-model of lipoprotein(a) was developed in the All of Us Research Program (data release 7), with the goal of predicting circulating lipoprotein(a) levels accurately across individuals of diverse genetic background. Briefly, the sequencing-based Lp(a) prediction model developed in UKB was used to predict circulating lipoprotein(a) levels attributable to each haplotype of each AoU participant (after re-optimizing the model using UKB whole-genome sequencing data). To develop the haplotype-based model, we identified single nucleotide polymorphism (SNP) haplotypes flanking the LPA locus on chromosome 6 shared by >20 individuals in the AoU cohort. Each haplotype, representing an *LPA* allele, was assigned the mean allele-specific Lp(a) concentration predicted for the haplotype’s carriers by the sequencing-based model. The standard deviation of these predicted Lp(a) concentrations was used as a measure of uncertainty. The LPA-haplotype model was then applied to phased genotype data imputed to the TOPMed reference panel from the PMBB, MGBB, and BioMe cohorts to predict circulating Lp(a) levels and identify individuals with elevated Lp(a) levels. A schematic example of the haplotype-model is presented in **eFigure 2** and **Figure 3**.

**Figure 2:**
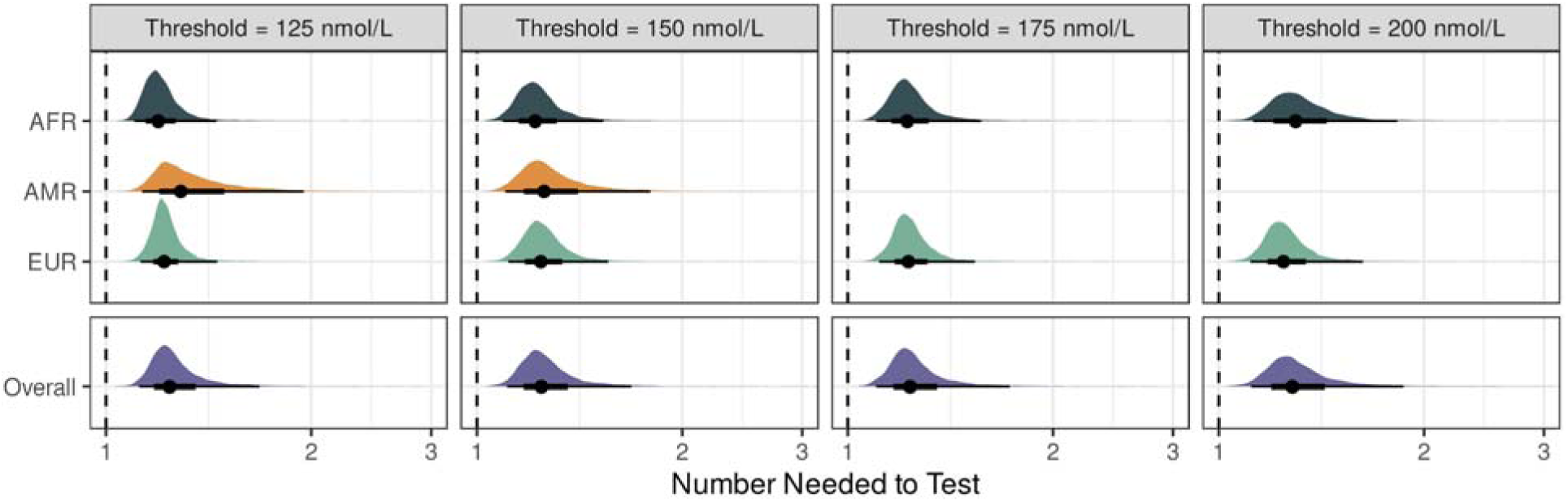
Number needed to test to identify 1 individual with an elevated lipoprotein(a) concentration above clinical thresholds. Number needed to test (NNT) represents the number of individuals predicted to have elevated Lp(a) who would require confirmatory clinical testing to identify one true elevation. Results shown across clinical thresholds (125, 150, 175, and 200 nmol/L) and stratified by genetically inferred ancestry. Distributions represent posterior estimates from Bayesian models; points and horizontal lines indicate posterior medians and 95% credible intervals. Dashed vertical line indicates NNT = 1 (perfect prediction). AFR = African; AMR = Admixed American; EUR = European.

**Figure 3:**
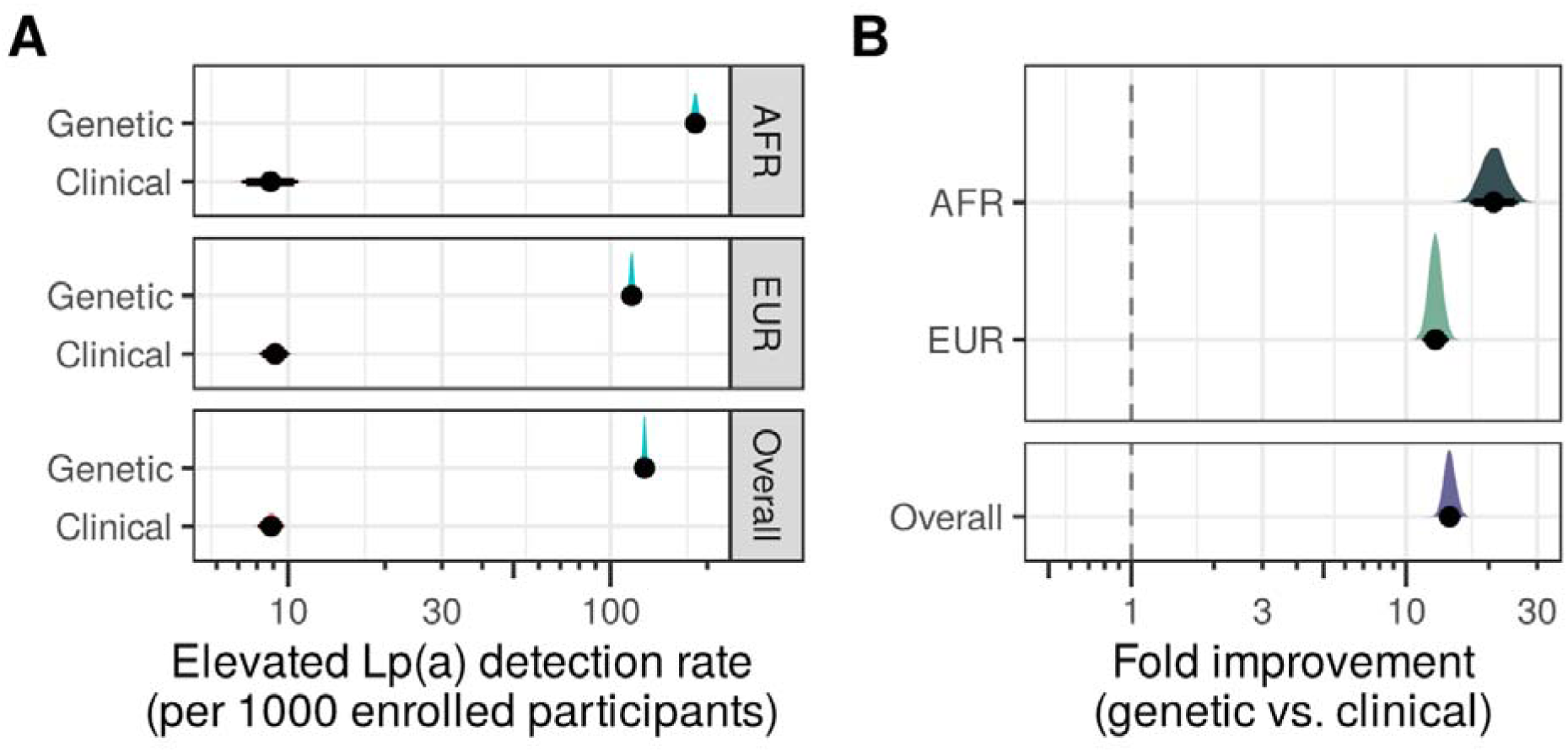
Rates of elevated lipoprotein(a) concentrations >125 nmol/L identified by haplotype-based genetic prediction versus clinical assessment. A) Detection rates of elevated Lp(a) (>125 nmol/L) per 1000 enrolled participants by clinical measurement versus genetic prediction (logDD scale). B) Fold improvement in detection rate comparing genetic prediction to clinical assessment; dashed vertical line indicates no improvement (fold change = 1). Distributions represent posterior estimates; points and horizontal lines indicate posterior medians and 95% credible intervals. AFR = African ancestry; EUR = European ancestry.

### Validation Cohorts

The Penn Medicine Biobank (PMBB) is a biobank of individuals receiving care at the University of Pennsylvania Health System. The PMBB cohort includes individuals with extensive clinical and genetic data, including genome-wide genotype data. Details of the cohort, as well as genotyping, imputation, and quality control have been previously described.^10^ The validation cohort included 1856 PMBB participants with available clinically-obtained Lp(a) measurements and genotype data collected through September 2025. PMBB participants were grouped into sub-populations based on imputed genotypes using pgsc_calc, which implements the Fast and Robust Ancestry Prediction by using Online singular value decomposition and Shrinkage Adjustment (FRAPOSA) method using the 1000 Genomes + Human Genome Diversity Panel reference population. Where necessary, clinically available Lp(a) measurements were converted from mg/dL to nmol/L using a conversion factor of 2.15.^11^

The Mass General Brigham Biobank (MGBB) is a biobank of individuals receiving care within the Mass General Brigham health system. Details of the cohort, as well as genotyping, imputation, and quality control have been previously described.^12^ The MGBB validation cohort included 1401 MGBB participants with available clinically-obtained Lp(a) measurements and genotype data collected through September 2025.

The Mount Sinai BioMe biobank is a biobank of individuals receiving care within the Mount Sinai health system. Details of the cohort, as well as genotyping, imputation, and quality control have been previously described.^13^ The BioMe validation cohort included 1686 BioMe participants with available clinically-obtained Lp(a) measurements and genotype data collected through September 2025.

### Performance of Existing Lp(a) PGS

Polygenic score weights for Lp(a) (EFO_0006925) were obtained from the PGS Catalog on September 28, 2025 and applied to the PMBB cohort to predict circulating Lp(a) levels. PGS weights were applied to PMBB genotypes using pgsc_calc.^14^ Because PGS performance may vary due to differences in genetic ancestry, the Z_norm2 method was applied, which normalizes score mean and variance using genetic principal components using the external 1000 Genomes reference population. The performance of existing Lp(a) PGS was assessed by comparing predicted and measured Lp(a) levels across population subgroups in the PMBB cohort. r-squared was used to quantify the performance of the polygenic scores in predicting continuous Lp(a) levels.

### Performance of the LPA-Haplotype Model

The LPA-haplotype model was applied externally to the PMBB, MGBB, and BioMe cohorts to predict circulating Lp(a) levels and identify individuals with elevated Lp(a) levels. For each individual, Lp(a) concentrations were generated corresponding to each LPA allele. The sum of these concentrations was used as the predicted Lp(a) level. Uncertainty measures of the predicted Lp(a) concentrations were also generated, with upper- and lower 95% confidence bounds derived from the standard error: 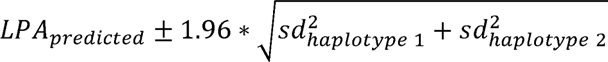 where *sd* is the standard deviation of allele-specific predicted Lp(a) measurements across AoU participants sharing a given haplotype. Predicted Lp(a) levels were compared to measured Lp(a) levels in the PMBB, MGBB, and BioMe cohorts. The performance of the LPA-haplotype model was assessed by comparing predicted and measured Lp(a) levels across genetically-defined population subgroups. The performance of the LPA-haplotype model was compared to existing PGS in the PMBB cohort. Across all cohorts, the ability of the LPA-haplotype model to identify individuals with elevated Lp(a) levels was evaluated by comparing predicted and measured Lp(a) levels in individuals with clinical elevations of Lp(a) (>125 nmol/L, as defined by AHA/ACC and NLA guidelines and international consensus statements).^3,15,16^ In sensitivity analyses, alternative Lp(a) thresholds of 150 nmol/L, 175 nmol/L, and 200 nmol/L were considered. Ancestry subgroups were included in ancestry-specific performance assessments when >30 individuals from each cohort above a given threshold were available. Performance measures used to assess the LPA-haplotype model included continuous r-squared, while sensitivity, specificity, positive predictive value, negative predictive value, positive- and negative- likelihood ratios, accuracy, and F1 score were used to identify individuals with elevated Lp(a) levels above each clinical threshold. The primary performance measures of interest were continuous r^2^ and PPV/number-needed-to-test.

### Statistical Analysis

Statistical analyses were performed using R version 4.3.2. Continuous variables are presented as mean ± standard deviation or median (interquartile range), while categorical variables are presented as counts and percentages. Performance of existing PGS and the LPA-haplotype model was assessed using boostrap resampling with 1000 iterations. Performance measures were combined across cohorts using a Bayesian mixed model including random effects for genetically-inferred ancestry and cohort. The difference in performance of the LPA-haplotype model and existing PGS in PMBB was quantified using a Bayesian analysis of variance.^17^ Number needed to test was calculated as the reciprocal of the positive predictive value. Rates of elevated Lp(a) > 125 nmol/L observed from current clinical assessment and haplotype-based genetic prediction, were estimated using Bayesian binomial models fit using the brms package in R.^18^ To account for imperfect prediction, the count of genetically-identified elevations was adjusted downward by the positive predictive value of the genetic prediction model.

## RESULTS

### Demographics of the Validation Cohorts

Among PMBB (n = 1856), MGBB (n = 1401), and BioMe (n = 1686) participants with available genotype and Lp(a) measurements, average age was 60 years, and 51% were female (**Table 1**). Rates of cardiovascular comorbidities among participants with clinically-obtained Lp(a) measurements were generally high (**eTable 1**). Consistent with prior reports, measured Lp(a) concentrations were significantly higher among AFR participants compared to those of other genetic backgrounds (Wilcoxon rank sum p < 0.01) (**eFigure 4**).

**Table 1:** Demographics of the PMBB, MGBB, and BioMe validation cohorts.

| Characteristic | PMBB (N=1,856) | MGBB (N=1,401) | BioMe (N=1,686) |
| --- | --- | --- | --- |
| Age (years) | 59 (49, 67) | 56 (47, 65) | 65 (56, 73) |
| Sex |  |  |  |
| Female | 841 (45%) | 687 (49%) | 976 (58%) |
| Male | 1,015 (55%) | 714 (51%) | 710 (42%) |
| Genetic ancestry |  |  |  |
| AFR | 230 (12%) | 67 (4.8%) | 614 (36%) |
| AMR | 47 (2.5%) | 74 (5.3%) | 174 (10%) |
| CSA | 41 (2.2%) |  | 81 (4.8%) |
| EAS | 23 (1.2%) | 28 (2.0%) | 65 (3.9%) |
| EUR | 1,515 (82%) | 1,210 (86%) | 648 (38%) |
| Other |  | 22 (1.6%) | 104 (6.2%) |
| Lp(a) (nmol/L) | 46 (17, 129) | 42 (16, 155) | 64 (27, 146) |

### Performance of Existing Lp(a) PGS

Performance of existing PGS for Lp(a) are presented in **eFigure 5** and **eTable 2**. Among the overall PMBB population, r^2^ ranged from 0.13 (95% CI 0.097 to 0.16) to 0.3 (95% CI 0.25 to 0.35). Within each score, we observed significant heterogenitiy in r^2^ across ancestries (Cochran’s Q p-value <0.001) (**eTable 3**). Consistent with prior reports, PGS performance among the AFR population was universally poor, with r^2^ ranging from 0.0042 (95% CI 4.3e-06 to 0.022) to 0.0093 (95% CI 2.5e-05 to 0.037). These results suggest existing PGS for Lp(a) have heterogenous performance across ancestries, with particularly poor performance among AFR individuals.

### Performance of the LPA-Haplotype Model in PMBB, MGBB, and BioMe

To address the poor performance of existing polygenic scores for Lp(a), we developed a haplotype-based model in the All of Us research program that assigns Lp(a) values based on haplotype sharing, an approach more robust to differences in genetic background. Predicted and observed concentrations of Lp(a) in the external PMBB, MGBB, and BioMe validation cohorts are presented in **Figure 1**. The overall r^2^ was 0.46 (95% CrI 0.32 to 0.6). Performance was consistent across cohorts and populations, ranging from 0.402 (95% CrI 0.201 to 0.571) among EAS to 0.53 (95% CrI 0.382 to 0.652) among EUR. The haplotype-based model substantially improved performance compared to existing polygenic scores with Δr² ranging from 0.200 (95% CrI: 0.199, 0.201) to 0.367 (95% CrI: 0.365, 0.368) (**eFigure 6**; **eTable 4**). The haplotype model also outperformed all existing PGS across the AFR, AMR, and EUR populations, with more variable performance among EAS and SAS populations likely due to small sample sizes (EAS n = 8; CSA n = 15) (see **eTable 2** and **eTable 3** for existing PGS performance).

Performance measures of the LPA-haplotype model in predicting elevated Lp(a) concentrations are presented in **eTable 5**. The overall positive predictive value for identifying individuals with elevated Lp(a) concentrations > 125 nmol/L was 0.81 (95% CrI 0.6 to 0.89), and across populations ranged from 0.777 (95% CrI 0.513 to 0.885) among AMR to 0.839 (95% CrI 0.689 to 0.909) among AFR. The overall positive likelihood ratio was 8.9 (95% CrI 2.9 to 17), and overall negative likelihood ratio was 0.52 (95% CrI 0.34 to 0.73). Results were similar when considering different thresholds for elevated Lp(a) concentrations. Full performance measures across populations are presented in **eTable 5**.

### Number Needed to Test

To quantify the case-finding utility of the haplotype model, the number needed to test (NNT) to identify 1 individual with an elevated lipoprotein(a) concentration >125 nmol/L based on their haplotype-predicted level was estimated (**Figure 2** and **eTable 6**). Overall, the NNT was 1.2 (95% CrI 1.1 to 1.7) and ranged from 1.2 (95% CrI 1.1 to 1.5) among AFR to 1.3 (95% CrI 1.1 to 1.9) among AMR. These results suggest that among individuals predicted to have elevated levels of Lp(a) based on existing genotype data, fewer than 1.5 individuals would need to undergo clinical testing to identify 1 individual with a true elevation >125 nmol/L.

### Rates of Elevated Lipoprotein(a) Concentrations by Genetics vs. Clinical Assessment

Despite cardiovascular guideline recommendations for universal one-time Lp(a) assessment among adults, rates of clinical Lp(a) assessment remain low. These low rates limit identification of individuals that may benefit from intensifying lipid-lowering treatment, for example with PCSK9 inhibitors, consideration of lipoprotein-apheresis for Lp(a)-lowering, family cascade screening, as well as identifying those who may benefit from upcoming Lp(a)-lowering therapies. To understand the potential role for genetic prediction of elevated Lp(a) to improve diagnosis rates among existing genotyped cohorts, we compared rates of elevated Lp(a) concentrations >125 nmol/L among PMBB participants who had undergone Lp(a) measurement as part of routine clinical care in comparison to estimated rates in the full genotyped cohort based on haplotype-predicted levels. The rate of identifying elevated Lp(a) was 128 per 1000 (95% CrI 125 to 130) among the genotyped cohort compared with 8.86 per 1000 (95% CrI 8.06 to 9.67) among those who had prior clinical Lp(a) measurement, corresponding to a 14.4-fold improvement (95% CrI 13.1 to 15.9; P(improvement) = 1) in detection rate. The highest rates of elevated Lp(a) identification using the haplotype model were among AFR participants: 184 per 1000 (95% CrI 176 to 191). These results indicate that when genotype information is available, identifying individuals with elevated Lp(a) concentrations based on genetics may offer an efficient and targeted approach for identifying at-risk patients when rates of clinical screening in current practice are low.

## DISCUSSION

In this study, we developed and validated a haplotype-based model for predicting lipoprotein(a) levels using genome-wide genotype data, demonstrating strong performance across diverse populations. As a highly heritable trait, genetic prediction of Lp(a) has been an exciting prospect for more than a decade, but has been hampered by complex methods, limited generalizability across diverse populations, and lack of widespread genetic testing.^1,5,7,19^ Here, we demonstrate that among 3 large biobank cohorts with existing genotype information, haplotype-based Lp(a) prediction has the potential to improve opportunistic identification of individuals with elevated Lp(a) concentrations. These results have important methodologic implications for the transferability of genetic predictions across diverse populations, and important clinical implications for efficiently identifying individuals who may be eligible for upcoming therapeutics and future clinical trials compared with current clinical practice.

Our findings demonstrate that haplotype-based Lp(a) prediction maintains consistent accuracy across genetically inferred ancestral groups, addressing a critical limitation of existing PGS approaches. Traditional PGS for Lp(a) show substantial performance degradation in non-European populations—particularly among individuals of African ancestry, where existing scores explain almost none of the variance in measured Lp(a) levels.^7^ This is especially concerning given that African-ancestry populations have higher median Lp(a) concentrations and higher prevalence of clinical elevations compared with European-ancestry populations.^11^ Rather than assuming that variant effects estimated in one population will transfer to others as in traditional PGS, our haplotype-based approach matches individuals by local chromosomal structure at the LPA locus, capturing the combined effects of repeat variation, rare coding variants, and common regulatory polymorphisms without requiring explicit modeling of each component. This haplotype-based framework is particularly well-suited to oligogenic traits with high heritability and limited polygenicity, like lipoprotein(a). The consistently strong performance of our model across ancestral groups—in contrast to the marked performance degradation seen with existing PGS—suggests that for appropriate traits, haplotype-based prediction can overcome the transferability limitations that have generated skepticism about clinical implementation of traditional PGS. As genetic data becomes available in additional populations, the haplotype reference panel can be readily extended. This characteristic may help address concerns about equitable implementation of genetic prediction tools across diverse populations.

Our results support an “opportunistic screening” strategy for elevated Lp(a) among individuals with existing genotype data, and raise the possibliity of more targeted testing strategies in the future as genotyping becomes more ubiquitous. The high positive predictive value of the haplotype model means that individuals predicted to have elevated Lp(a) can be efficiently triaged for confirmatory clinical testing, with a number needed to test of 1.2 (95% CrI 1.1 to 1.7) to identify one true elevation. This approach could be particularly valuable in biobank settings, where large numbers of individuals have genotype data available but few have undergone Lp(a) measurement (<1% in our validation cohorts). By leveraging existing genetic data, healthcare systems could identify individuals at elevated cardiovascular risk due to high Lp(a) concentrations without the need for universal biochemical screening. Although the negative predictive value of 0.79 (95% CrI 0.56 to 0.9) and negative likelihood ratio of 0.52 (95% CrI 0.34 to 0.73) may not be sufficient to exclude clinically-significant elevations in Lp(a), the overall positive likelihood ratio of 8.9 (95% CrI 2.9 to 17) at a threshold of 125 nmol/L and 11 (95% CrI 3.3 to 23) at a threshold of 150 nmol/L approaches/exceeds the diagnostic threshold of 10, indicating the potential to rule-in clinically significant elevations via genetic testing.^20^ Prior studies have demonstrated that opportunistic genetic screening offers the potential to improve management of other inherited dyslipidemias like familial hypercholesterolemia.^21^ Given the large proportion of the population estimated to have genetically-elevated Lp(a) levels, opportunistic genetic screening has the potential to address gaps in clinical Lp(a) assessment. This opportunistic strategy is complementary to existing clinical prediction models for elevated Lp(a), which have been developed using routinely available clinical variables, but demonstrate only modest discriminative ability.^22^ Because Lp(a) levels are 70–90% genetically determined, clinical models are inherently limited when genetic information is available. Clinical models may be useful for triaging patients toward Lp(a) measurement in routine practice, while genetic prediction may enable opportunistic screening among already-genotyped populations including biobanks and individuals pursing direct-to-consumer genetic testing for family or ancestry information. As population-based genotyping becomes more common, accurate genetic predictions models of Lp(a) may enable a future where confirmatory testing and clinical follow-up can be performed more selectively.

The clinical relevance of identifying elevated Lp(a) is increasing rapidly. Multiple Lp(a)-lowering therapies are in late-stage clinical trials, including pelacarsen (an antisense oligonucleotide in the Lp(a)HORIZON trial), olpasiran (a small interfering RNA in the OCEAN(a)-Outcomes trial), lepodisiarn (a small interfering RNA in the ACCLAIM-lipoprotein(a) trial), and muvalaplin (an oral small molecule in the MOVE-Lp(a) trial).^23^ If these therapies demonstrate clinical benefit, demand for Lp(a) testing will increase substantially, yet the historical pace of changing clinical practice suggests the current testing gap may persist. In the near term, opportunistic genetic screening could facilitate enrollment in ongoing and future clinical trials by identifying eligible individuals among existing biobank populations. For the tens of millions individuals who have already undergone genotyping for other purposes, and the growth of genotyping more broadly, Lp(a) prediction may represent a valuable secondary use of existing data.

Our findings should be interpreted in the context of several limitations. First, although our validation cohorts included participants of African, Admixed American, European, Central/South Asian, and East Asian genetic ancestry, these groups do not fully capture global human genetic diversity. Performance in underrepresented populations—including Indigenous populations and those with complex admixture patterns—remains to be established. However, the haplotype-based framework can be readily extended as genetic data from additional populations becomes available. Second, validation cohorts were derived from academic medical center biobanks, which may not be fully representative of the general population, although the distribution of Lp(a) mirrored those expected from the general population.^11^ Because Lp(a) levels are predominantly genetically determined with minimal influence from clinical or environmental factors, the model’s performance is unlikely to differ substantially in broader populations. Notably, prior clinical prediction models for Lp(a) based on demographic and laboratory variables have demonstrated substantially lower performance, underscoring the dominant role of genetic factors.^22^ Although we also benchmarked performance against existing PGS for Lp(a) from the PGS Catalog, other PGS-based models have also been developed and evaluated with mixed performance.^8,19,24^ Third, while elevated Lp(a) has been robustly linked to atherosclerotic cardiovascular disease and aortic stenosis in epidemiologic and genetic studies, we did not assess clinical outcomes.^6,25^ The impact of genetically-predicted Lp(a) elevation on cardiovascular events—and the clinical utility of early identification—remains to be established in prospective studies. Finally, our haplotype model predicts Lp(a) concentrations based on genetic data alone and does not incorporate clinical variables that may modestly influence levels, such as kidney function or inflammatory states. For clinical decision-making, confirmatory laboratory measurement of Lp(a) remains appropriate for individuals identified as likely elevated by genetic prediction.

## Conclusions

Haplotype-based genetic prediction of Lp(a) demonstrates strong, consistent performance across diverse populations using existing genotype data. This approach enables opportunistic screening for elevated Lp(a) among the tens of millions of individuals who have already undergone genetic testing, offering a potential strategy to close the current gap in clinical Lp(a) assessment.

## ACKNOWLEDGEMENTS

We acknowledge the Penn Medicine BioBank (PMBB) for providing data and thank the patient-participants of Penn Medicine who consented to participate in this research program. We would also like to thank the Penn Medicine BioBank team and Regeneron Genetics Center for providing genetic variant data for analysis. The PMBB is approved under IRB protocol# 813913 and supported by Perelman School of Medicine at University of Pennsylvania, a gift from the Smilow family, and the National Center for Advancing Translational Sciences of the National Institutes of Health under CTSA award number UL1TR001878.

The All of Us Research Program is supported by the National Institutes of Health, Office of the Director: Regional Medical Centers: 1 OT2 OD026549; 1 OT2 OD026554; 1 OT2 OD026557; 1 OT2 OD026556; 1 OT2 OD026550; 1 OT2 OD 026552; 1 OT2 OD026553; 1 OT2 OD026548; 1 OT2 OD026551; 1 OT2 OD026555; IAA #: AOD 16037; Federally Qualified Health Centers: HHSN 263201600085U; Data and Research Center: 5 U2C OD023196; Biobank: 1 U24 OD023121; The Participant Center: U24 OD023176; Participant Technology Systems Center: 1 U24 OD023163; Communications and Engagement: 3 OT2 OD023205; 3 OT2 OD023206; and Community Partners: 1 OT2 OD025277; 3 OT2 OD025315; 1 OT2 OD025337; 1 OT2 OD025276. In addition, the All of Us Research Program would not be possible without the partnership of its participants.

The contributions of the NIH author(s) are considered Works of the United States Government. The findings and conclusions presented in this paper are those of the author(s) and do not necessarily reflect the views of the NIH or the U.S. Department of Health and Human Services.

## DATA AVAILABILITY

The haplotype-based Lp(a) prediction model will be made available through a public repository upon publication. The Penn Medicine BioBank (PMBB) data utilized in this study can be accessed by qualified researchers through an application process detailed at https://pmbb.med.upenn.edu. The Mass General Brigham Biobank can be contacted at. The Mt. Sinai BioMe biobank can be contacted at. The All of Us Research Program data are accessible to registered researchers via the Researcher Workbench at https://www.researchallofus.org.

---

**eFigure 1:**
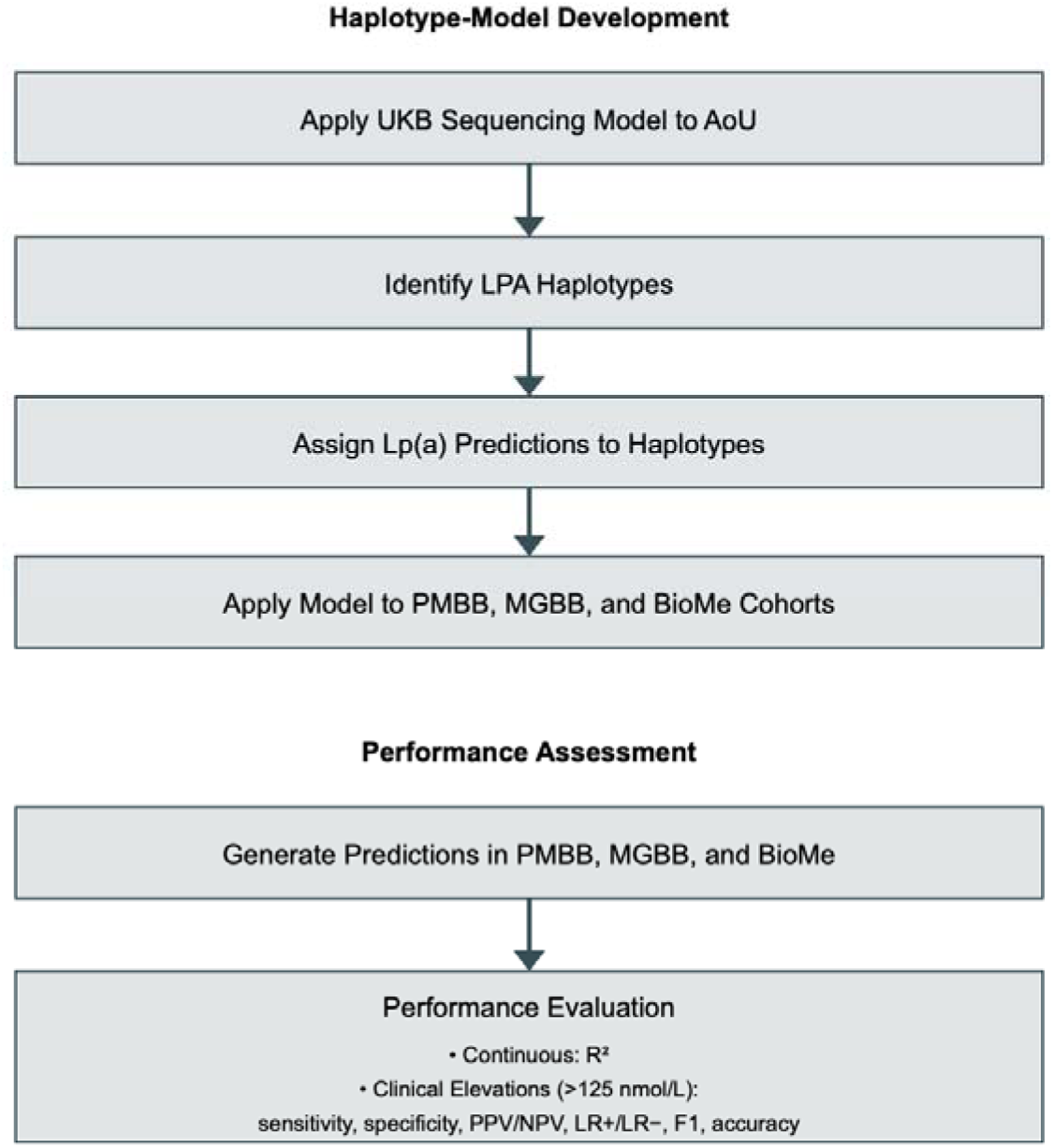
Schematic representation of the study design including haplotype-model development in the All of Us Research Program and external validation in PMBB, MGBB, and BioMe cohorts.

**eFigure 2:**
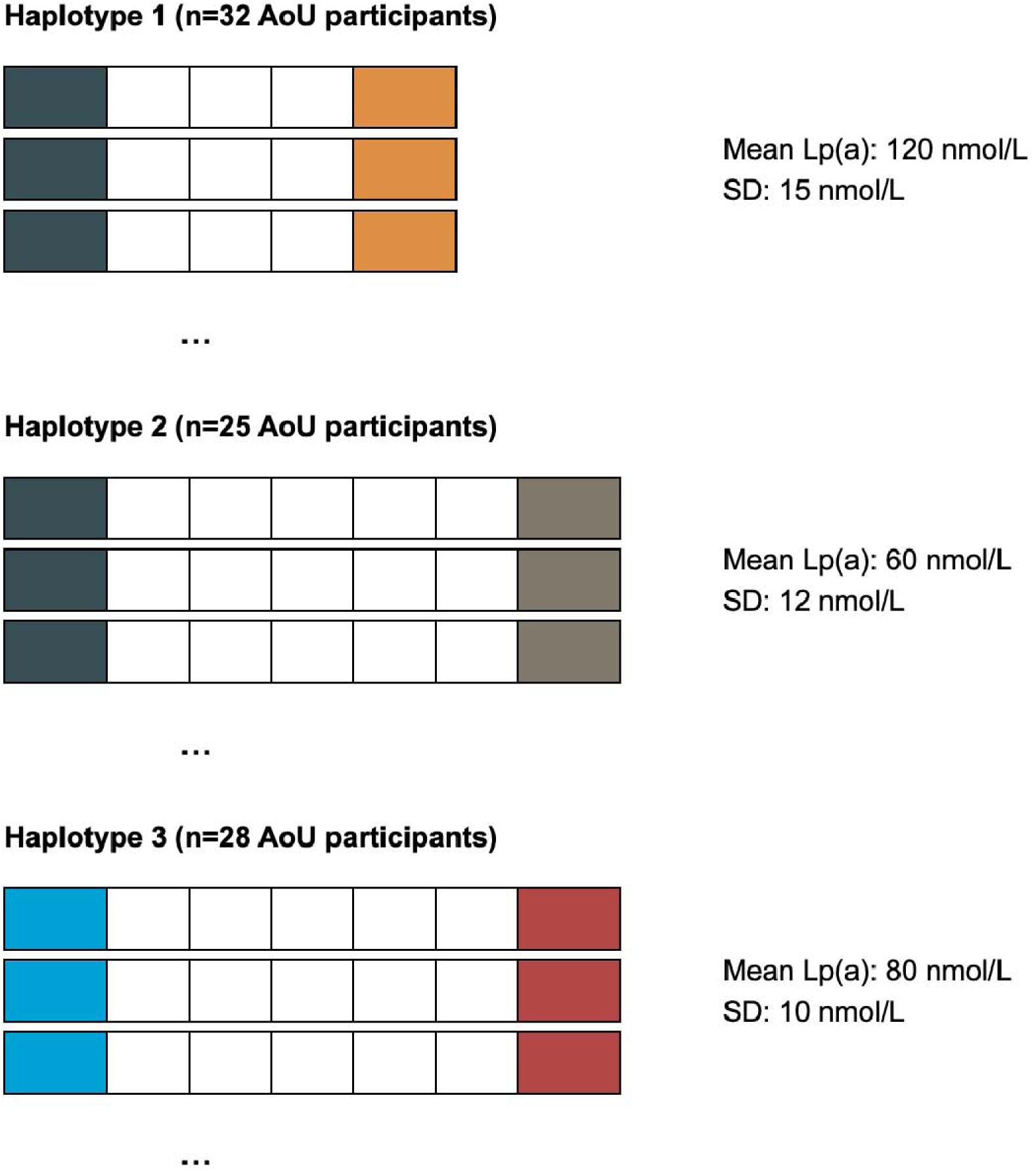
In the All of Us Research Program, participants were grouped by haplotypes flanking the LPA locus. Each haplotype was assigned the Lp(a) concentration from a sequencing-based model previously developed in the UK Biobank, with standard deviation used as a measure of uncertainty.

**eFigure 3:**
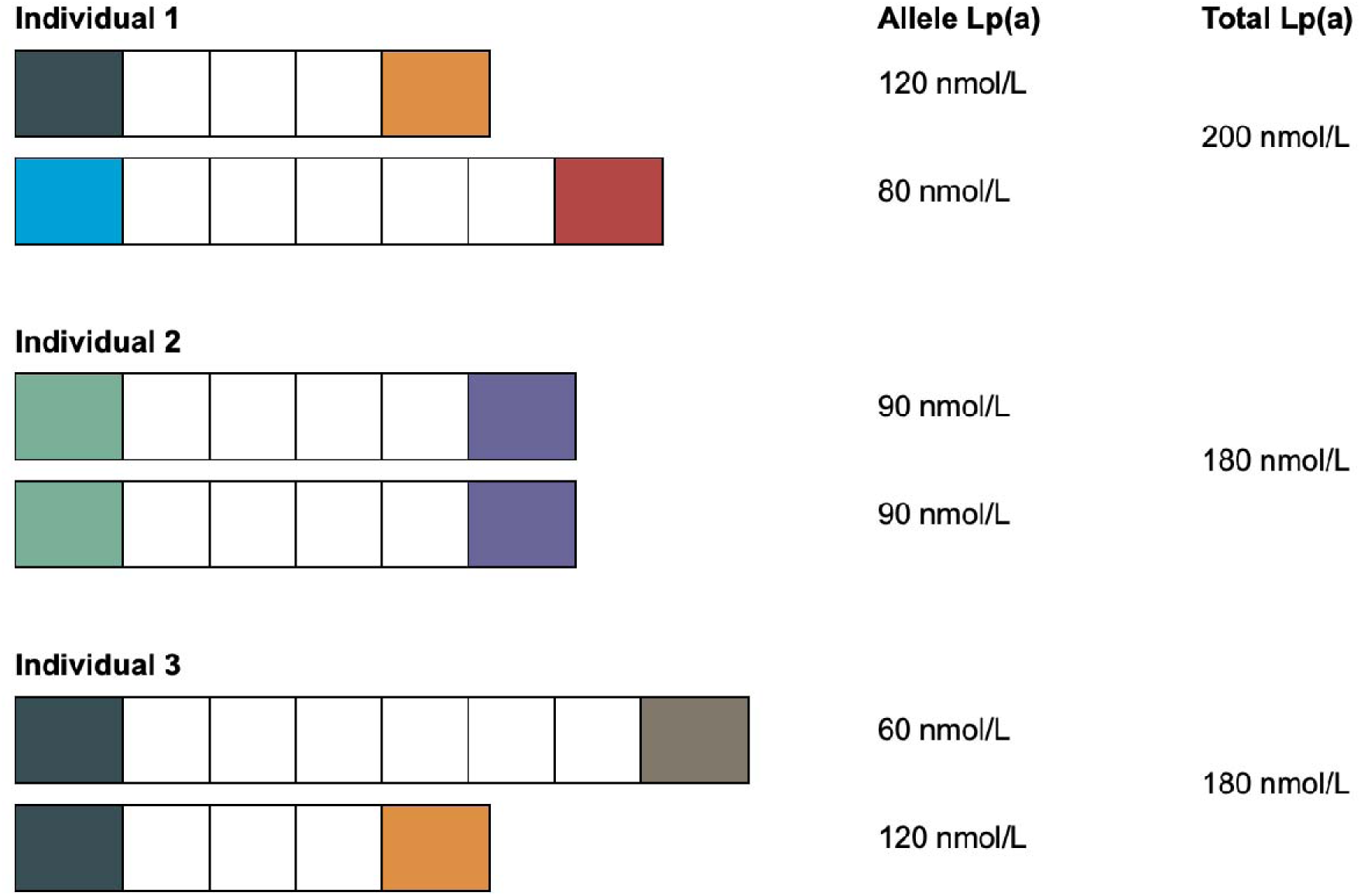
For prediction in external cohorts, individuals were matched to reference haplotypes based on shared chromosomal segments. Total predicted Lp(a) was calculated as the sum of predictions from each allele.

**eFigure 4:**
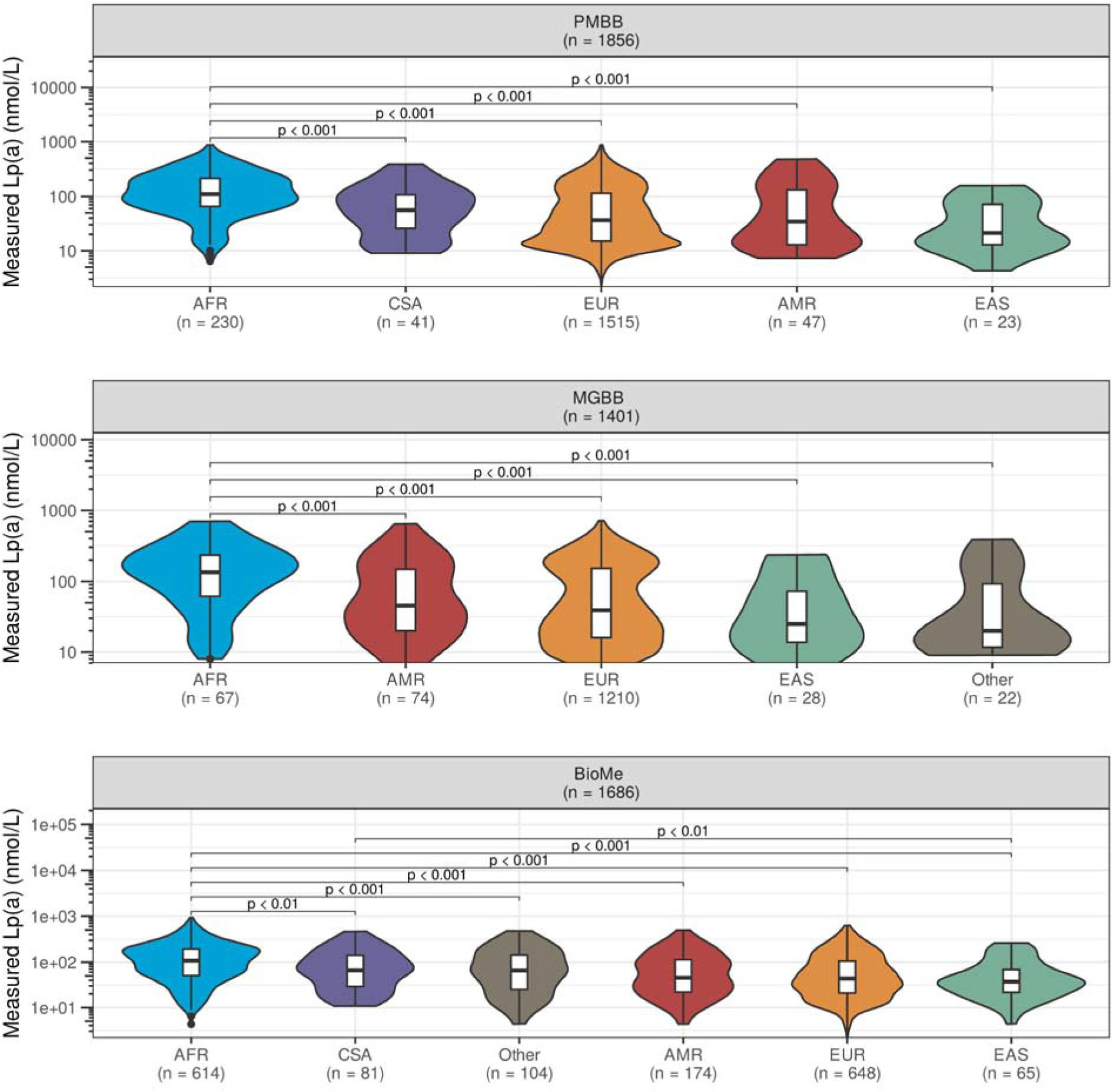
Lp(a) Concentrations Across Population Subgroups. Violin plots showing the distribution of clinically measured Lp(a) concentrations (logDD scale) stratified by genetically inferred ancestry in PMBB, MGBB, and BioMe cohorts. Boxplots indicate median and interquartile range. P-values from Wilcoxon rank-sum tests. Sample sizes shown in parentheses. AFR = African; AMR = Admixed American; CSA = Central/South Asian; EAS = East Asian; EUR = European.

**eFigure 5:**
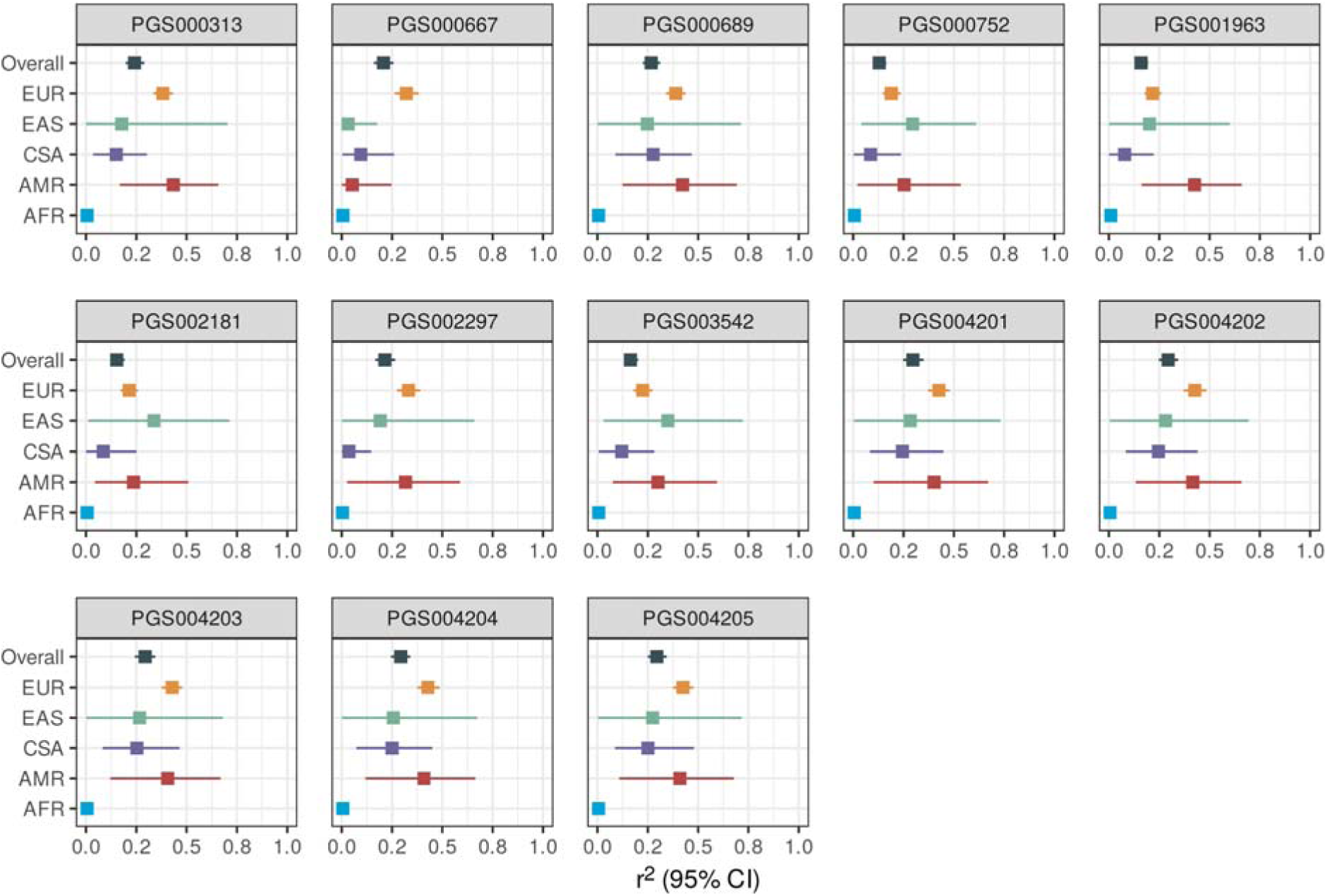
Performance of Existing Lp(a) Polygenic Scores in the Penn Medicine Biobank. R² (variance explained) with 95% confidence intervals for 13 published polygenic scores for Lp(a) obtained from the PGS Catalog, stratified by genetically inferred ancestry. Scores were applied using pgsc_calc with Z_norm2 ancestry adjustment. AFR = African; AMR = Admixed American; CSA = Central/South Asian; EAS = East Asian; EUR = European.

**eFigure 6:**
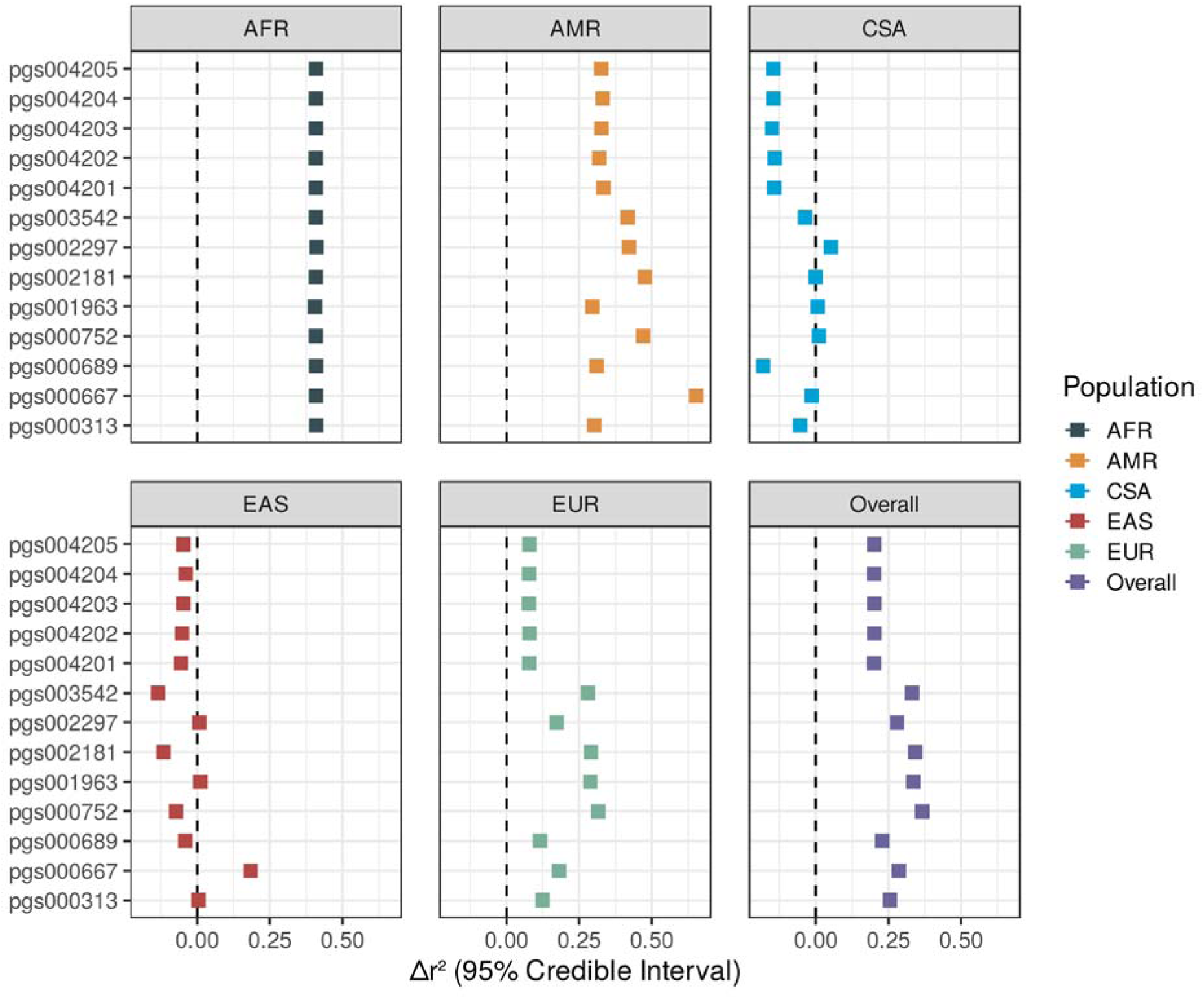
Improvement in prediction performance of the LPA-haplotype model compared with existing polygenic scores. Δr² (difference in variance explained) between the haplotype model and each of 13 published polygenic scores, stratified by genetically inferred ancestry. Positive values indicate superior performance of the haplotype model. Points and horizontal lines indicate posterior medians and 95% credible intervals from Bayesian analysis of variance. Note consistent improvement across all scores in AFR, AMR, and EUR populations. AFR = African; AMR = Admixed American; CSA = Central/South Asian; EAS = East Asian; EUR = European.

**eTable 1:** Clinical Characteristics of Validation Cohorts.

| Characteristic | PMBB (N=1,856) | MGBB (N=1,401) | BioMe (N=1,686) |
| --- | --- | --- | --- |
| Age (years) | 59 (49, 67) | 56 (47, 65) | 65 (56, 73) |
| Sex |  |  |  |
| Female | 841 (45%) | 687 (49%) | 976 (58%) |
| Male | 1,015 (55%) | 714 (51%) | 710 (42%) |
| Ischemic heart disease | 469 (25%) |  | 723 (43%) |
| Hypertension | 844 (45%) |  | 1,260 (75%) |
| Hyperlipidemia | 1,435 (77%) |  |  |
| Heart failure | 151 (8.1%) |  | 297 (18%) |
| Chronic kidney disease | 224 (12%) |  | 530 (31%) |
| Diabetes | 299 (16%) |  | 663 (39%) |
| Atrial fibrillation | 189 (10%) |  | 178 (11%) |
| Peripheral vascular disease | 97 (5.2%) |  | 391 (23%) |
| Ischemic stroke | 48 (2.6%) |  | 138 (8.2%) |

**eTable 2:** Performance of Existing Lp(a) Polygenic Scores.

| MostSimilarPop | PGS | .metric | estimate | sd | lower_ci | upper_ci |
| --- | --- | --- | --- | --- | --- | --- |
| AFR | PGS000313 | rsq | 0.00529 | 0.00659 | 0.00000934 | 0.0218 |
| AFR | PGS000667 | rsq | 0.00577 | 0.00725 | 0.00000910 | 0.0256 |
| AFR | PGS000689 | rsq | 0.00513 | 0.00683 | 0.00000485 | 0.0237 |
| AFR | PGS000752 | rsq | 0.00642 | 0.00786 | 0.00000526 | 0.0286 |
| AFR | PGS001963 | rsq | 0.00929 | 0.0102 | 0.0000247 | 0.0367 |
| AFR | PGS002181 | rsq | 0.00576 | 0.00756 | 0.00000624 | 0.0274 |
| AFR | PGS002297 | rsq | 0.00422 | 0.00574 | 0.00000432 | 0.0219 |
| AFR | PGS003542 | rsq | 0.00596 | 0.00787 | 0.00000524 | 0.0288 |
| AFR | PGS004201 | rsq | 0.00512 | 0.00734 | 0.00000421 | 0.0268 |
| AFR | PGS004202 | rsq | 0.00534 | 0.00686 | 0.00000707 | 0.0250 |
| AFR | PGS004203 | rsq | 0.00536 | 0.00710 | 0.00000320 | 0.0261 |
| AFR | PGS004204 | rsq | 0.00535 | 0.00736 | 0.00000547 | 0.0245 |
| AFR | PGS004205 | rsq | 0.00565 | 0.00760 | 0.00000649 | 0.0248 |
| AMR | PGS000313 | rsq | 0.434 | 0.126 | 0.168 | 0.658 |
| AMR | PGS000667 | rsq | 0.0525 | 0.0725 | 0.0000724 | 0.248 |
| AMR | PGS000689 | rsq | 0.422 | 0.146 | 0.124 | 0.693 |
| AMR | PGS000752 | rsq | 0.253 | 0.138 | 0.0212 | 0.534 |
| AMR | PGS001963 | rsq | 0.425 | 0.129 | 0.162 | 0.661 |
| AMR | PGS002181 | rsq | 0.236 | 0.122 | 0.0450 | 0.509 |
| AMR | PGS002297 | rsq | 0.317 | 0.153 | 0.0267 | 0.588 |
| AMR | PGS003542 | rsq | 0.301 | 0.136 | 0.0757 | 0.594 |
| AMR | PGS004201 | rsq | 0.401 | 0.152 | 0.101 | 0.672 |
| AMR | PGS004202 | rsq | 0.416 | 0.141 | 0.133 | 0.660 |
| AMR | PGS004203 | rsq | 0.407 | 0.143 | 0.120 | 0.669 |
| AMR | PGS004204 | rsq | 0.408 | 0.146 | 0.118 | 0.664 |
| AMR | PGS004205 | rsq | 0.409 | 0.147 | 0.108 | 0.678 |
| CSA | PGS000313 | rsq | 0.150 | 0.0703 | 0.0346 | 0.302 |
| CSA | PGS000667 | rsq | 0.0945 | 0.0692 | 0.00316 | 0.260 |
| CSA | PGS000689 | rsq | 0.276 | 0.0985 | 0.0887 | 0.468 |
| CSA | PGS000752 | rsq | 0.0863 | 0.0609 | 0.00276 | 0.238 |
| CSA | PGS001963 | rsq | 0.0782 | 0.0613 | 0.000359 | 0.223 |
| CSA | PGS002181 | rsq | 0.0857 | 0.0688 | 0.000472 | 0.250 |
| CSA | PGS002297 | rsq | 0.0356 | 0.0419 | 0.0000289 | 0.147 |
| CSA | PGS003542 | rsq | 0.121 | 0.0714 | 0.00650 | 0.282 |
| CSA | PGS004201 | rsq | 0.244 | 0.0981 | 0.0837 | 0.448 |
| CSA | PGS004202 | rsq | 0.246 | 0.0961 | 0.0842 | 0.441 |
| CSA | PGS004203 | rsq | 0.253 | 0.0976 | 0.0817 | 0.465 |
| CSA | PGS004204 | rsq | 0.250 | 0.101 | 0.0725 | 0.451 |
| CSA | PGS004205 | rsq | 0.251 | 0.101 | 0.0868 | 0.479 |
| EAS | PGS000313 | rsq | 0.177 | 0.193 | 0.000263 | 0.704 |
| EAS | PGS000667 | rsq | 0.0319 | 0.0541 | 0.0000371 | 0.176 |
| EAS | PGS000689 | rsq | 0.248 | 0.206 | 0.000527 | 0.713 |
| EAS | PGS000752 | rsq | 0.295 | 0.155 | 0.0407 | 0.611 |
| EAS | PGS001963 | rsq | 0.201 | 0.166 | 0.000583 | 0.600 |
| EAS | PGS002181 | rsq | 0.337 | 0.193 | 0.0104 | 0.714 |
| EAS | PGS002297 | rsq | 0.191 | 0.185 | 0.000416 | 0.658 |
| EAS | PGS003542 | rsq | 0.349 | 0.189 | 0.0305 | 0.723 |
| EAS | PGS004201 | rsq | 0.283 | 0.202 | 0.00275 | 0.731 |
| EAS | PGS004202 | rsq | 0.281 | 0.196 | 0.00399 | 0.694 |
| EAS | PGS004203 | rsq | 0.266 | 0.197 | 0.00116 | 0.680 |
| EAS | PGS004204 | rsq | 0.256 | 0.194 | 0.000801 | 0.673 |
| EAS | PGS004205 | rsq | 0.274 | 0.200 | 0.00233 | 0.717 |
| EUR | PGS000313 | rsq | 0.382 | 0.0245 | 0.333 | 0.433 |
| EUR | PGS000667 | rsq | 0.321 | 0.0308 | 0.262 | 0.382 |
| EUR | PGS000689 | rsq | 0.388 | 0.0256 | 0.340 | 0.438 |
| EUR | PGS000752 | rsq | 0.189 | 0.0224 | 0.147 | 0.236 |
| EUR | PGS001963 | rsq | 0.217 | 0.0215 | 0.177 | 0.260 |
| EUR | PGS002181 | rsq | 0.214 | 0.0227 | 0.171 | 0.259 |
| EUR | PGS002297 | rsq | 0.331 | 0.0294 | 0.274 | 0.390 |
| EUR | PGS003542 | rsq | 0.225 | 0.0241 | 0.178 | 0.273 |
| EUR | PGS004201 | rsq | 0.426 | 0.0274 | 0.372 | 0.480 |
| EUR | PGS004202 | rsq | 0.426 | 0.0283 | 0.370 | 0.484 |
| EUR | PGS004203 | rsq | 0.429 | 0.0273 | 0.376 | 0.479 |
| EUR | PGS004204 | rsq | 0.428 | 0.0278 | 0.375 | 0.487 |
| EUR | PGS004205 | rsq | 0.425 | 0.0257 | 0.375 | 0.476 |
| Overall | PGS000313 | rsq | 0.241 | 0.0238 | 0.197 | 0.290 |
| Overall | PGS000667 | rsq | 0.208 | 0.0248 | 0.159 | 0.257 |
| Overall | PGS000689 | rsq | 0.267 | 0.0223 | 0.225 | 0.312 |
| Overall | PGS000752 | rsq | 0.129 | 0.0174 | 0.0975 | 0.164 |
| Overall | PGS001963 | rsq | 0.159 | 0.0172 | 0.128 | 0.194 |
| Overall | PGS002181 | rsq | 0.153 | 0.0183 | 0.121 | 0.192 |
| Overall | PGS002297 | rsq | 0.214 | 0.0266 | 0.166 | 0.265 |
| Overall | PGS003542 | rsq | 0.164 | 0.0182 | 0.131 | 0.202 |
| Overall | PGS004201 | rsq | 0.296 | 0.0253 | 0.247 | 0.350 |
| Overall | PGS004202 | rsq | 0.294 | 0.0249 | 0.248 | 0.344 |
| Overall | PGS004203 | rsq | 0.293 | 0.0254 | 0.243 | 0.344 |
| Overall | PGS004204 | rsq | 0.293 | 0.0257 | 0.244 | 0.341 |
| Overall | PGS004205 | rsq | 0.295 | 0.0244 | 0.249 | 0.344 |

**eTable 3:**
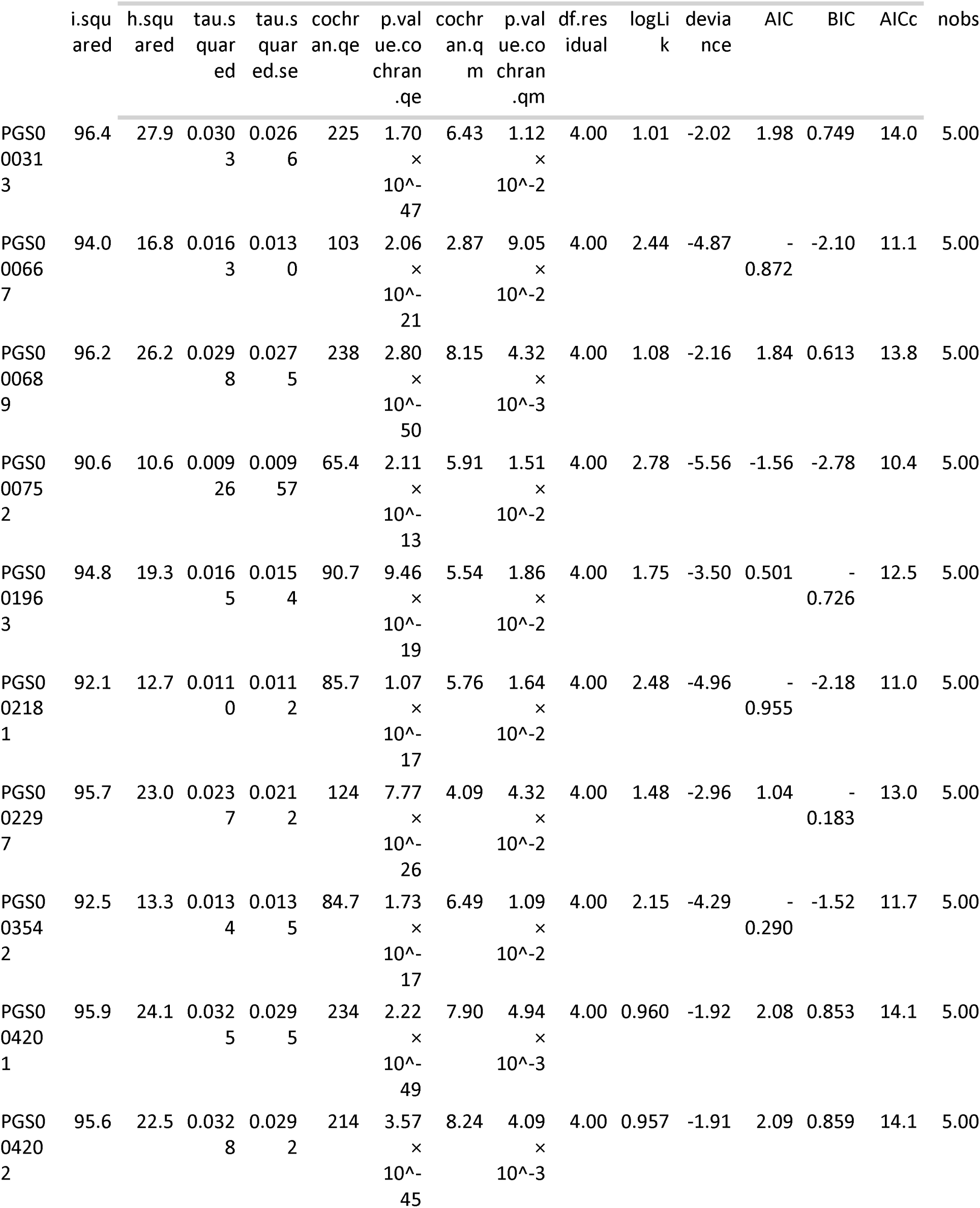
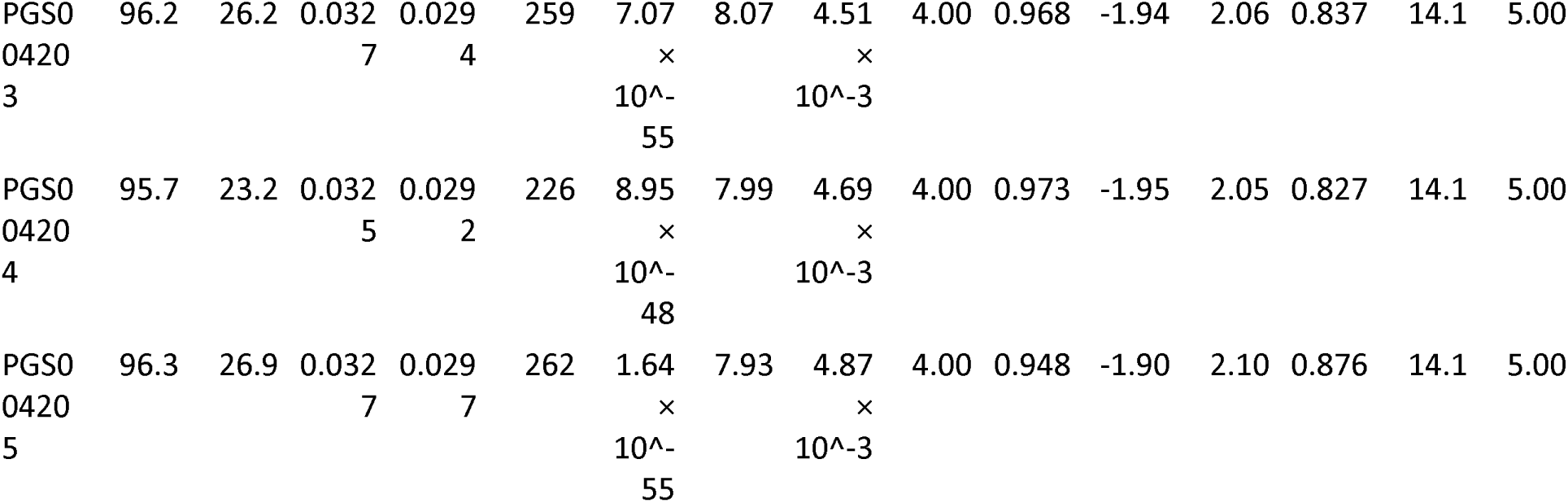
r^2^ Heterogeneity of Existing Lp(a) Polygenic Scores Across Ancestries.

**eTable 4:** Performance improvement of the LPA-haplotype model vs. existing PGS for Lp(a)

| group | contrast | probability | mean | lower | upper | pract_ne<br>g | pract_eq<br>uiv | pract_pos | n_obs |
| --- | --- | --- | --- | --- | --- | --- | --- | --- | --- |
| Overall | haplotype vs<br>pgs000313_hm_pos_gr_ch38 | 1.00 | 0.255 | 0.254 | 0.257 | 0 | 0 | 1.00 | 683 |
| Overall | haplotype vs<br>pgs000667_hm_pos_gr_ch38 | 1.00 | 0.286 | 0.285 | 0.288 | 0 | 0 | 1.00 | 683 |
| Overall | haplotype vs<br>pgs000689_hm_pos_gr_ch38 | 1.00 | 0.228 | 0.227 | 0.229 | 0 | 0 | 1.00 | 683 |
| Overall | haplotype vs<br>pgs000752_hm_pos_gr_ch38 | 1.00 | 0.367 | 0.365 | 0.368 | 0 | 0 | 1.00 | 683 |
| Overall | haplotype vs<br>pgs001963_hm_pos_gr_ch38 | 1.00 | 0.336 | 0.334 | 0.337 | 0 | 0 | 1.00 | 683 |
| Overall | haplotype vs<br>pgs002181_hm_pos_gr_ch38 | 1.00 | 0.342 | 0.341 | 0.344 | 0 | 0 | 1.00 | 683 |
| Overall | haplotype vs<br>pgs002297_hm_pos_gr_ch38 | 1.00 | 0.279 | 0.278 | 0.281 | 0 | 0 | 1.00 | 683 |
| Overall | haplotyp<br>e vs<br>pgs00354<br>2_hm_po<br>s_gr_ch3<br>8 | 1.00 | 0.332 | 0.330 | 0.333 | 0 | 0 | 1.00 | 683 |
| Overall | haplotyp<br>e vs<br>pgs00420<br>1_hm_po<br>s_gr_ch3<br>8 | 1.00 | 0.200 | 0.199 | 0.201 | 0 | 0 | 1.00 | 683 |
| Overall | haplotyp<br>e vs<br>pgs00420<br>2_hm_po<br>s_gr_ch3<br>8 | 1.00 | 0.201 | 0.200 | 0.203 | 0 | 0 | 1.00 | 683 |
| Overall | haplotyp<br>e vs<br>pgs00420<br>3_hm_po<br>s_gr_ch3<br>8 | 1.00 | 0.201 | 0.200 | 0.202 | 0 | 0 | 1.00 | 683 |
| Overall | haplotyp<br>e vs<br>pgs00420<br>4_hm_po<br>s_gr_ch3<br>8 | 1.00 | 0.200 | 0.199 | 0.202 | 0 | 0 | 1.00 | 683 |
| Overall | haplotyp<br>e vs<br>pgs00420<br>5_hm_po<br>s_gr_ch3<br>8 | 1.00 | 0.201 | 0.200 | 0.202 | 0 | 0 | 1.00 | 683 |
| AFR | haplotyp<br>e vs<br>pgs00031<br>3_hm_po<br>s_gr_ch3<br>8 | 1.00 | 0.409 | 0.408 | 0.411 | 0 | 0 | 1.00 | 84.0 |
| AFR | haplotyp<br>e vs<br>pgs00066<br>7_hm_po<br>s_gr_ch3<br>8 | 1.00 | 0.409 | 0.407 | 0.411 | 0 | 0 | 1.00 | 84.0 |
| AFR | haplotyp<br>e vs<br>pgs00068<br>9_hm_po<br>s_gr_ch3<br>8 | 1.00 | 0.409 | 0.407 | 0.411 | 0 | 0 | 1.00 | 84.0 |
| AFR | haplotyp<br>e vs<br>pgs00075<br>2_hm_po<br>s_gr_ch3<br>8 | 1.00 | 0.408 | 0.406 | 0.410 | 0 | 0 | 1.00 | 84.0 |
| AFR | haplotyp<br>e vs<br>pgs00196<br>3_hm_po<br>s_gr_ch3<br>8 | 1.00 | 0.405 | 0.403 | 0.407 | 0 | 0 | 1.00 | 84.0 |
| AFR | haplotyp<br>e vs<br>pgs00218<br>1_hm_po<br>s_gr_ch3<br>8 | 1.00 | 0.408 | 0.406 | 0.410 | 0 | 0 | 1.00 | 84.0 |
| AFR | haplotyp<br>e vs<br>pgs00229<br>7_hm_po<br>s_gr_ch3<br>8 | 1.00 | 0.411 | 0.409 | 0.413 | 0 | 0 | 1.00 | 84.0 |
| AFR | haplotyp<br>e vs<br>pgs00354<br>2_hm_po<br>s_gr_ch3<br>8 | 1.00 | 0.408 | 0.406 | 0.410 | 0 | 0 | 1.00 | 84.0 |
| AFR | haplotyp<br>e vs<br>pgs00420<br>1_hm_po<br>s_gr_ch3<br>8 | 1.00 | 0.408 | 0.406 | 0.410 | 0 | 0 | 1.00 | 84.0 |
| AFR | haplotyp<br>e vs<br>pgs00420<br>2_hm_po<br>s_gr_ch3<br>8 | 1.00 | 0.408 | 0.406 | 0.410 | 0 | 0 | 1.00 | 84.0 |
| AFR | haplotyp<br>e vs<br>pgs00420<br>3_hm_po<br>s_gr_ch3<br>8 | 1.00 | 0.408 | 0.407 | 0.410 | 0 | 0 | 1.00 | 84.0 |
| AFR | haplotyp<br>e vs<br>pgs00420<br>4_hm_po<br>s_gr_ch3<br>8 | 1.00 | 0.409 | 0.407 | 0.410 | 0 | 0 | 1.00 | 84.0 |
| AFR | haplotyp<br>e vs<br>pgs00420<br>5_hm_po<br>s_gr_ch3<br>8 | 1.00 | 0.409 | 0.407 | 0.410 | 0 | 0 | 1.00 | 84.0 |
| AMR | haplotyp<br>e vs<br>pgs00031<br>3_hm_po<br>s_gr_ch3<br>8 | 1.00 | 0.302 | 0.294 | 0.311 | 0 | 0 | 1.00 | 17.0 |
| AMR | haplotyp<br>e vs<br>pgs00066<br>7_hm_po<br>s_gr_ch3<br>8 | 1.00 | 0.653 | 0.644 | 0.662 | 0 | 0 | 1.00 | 17.0 |
| AMR | haplotyp<br>e vs<br>pgs00068<br>9_hm_po<br>s_gr_ch3<br>8 | 1.00 | 0.310 | 0.301 | 0.319 | 0 | 0 | 1.00 | 17.0 |
| AMR | haplotyp<br>e vs<br>pgs00075<br>2_hm_po<br>s_gr_ch3<br>8 | 1.00 | 0.470 | 0.461 | 0.478 | 0 | 0 | 1.00 | 17.0 |
| AMR | haplotyp<br>e vs<br>pgs00196<br>3_hm_po<br>s_gr_ch3<br>8 | 1.00 | 0.296 | 0.287 | 0.304 | 0 | 0 | 1.00 | 17.0 |
| AMR | haplotyp<br>e vs<br>pgs00218<br>1_hm_po<br>s_gr_ch3<br>8 | 1.00 | 0.477 | 0.468 | 0.485 | 0 | 0 | 1.00 | 17.0 |
| AMR | haplotyp<br>e vs<br>pgs00229<br>7_hm_po<br>s_gr_ch3<br>8 | 1.00 | 0.422 | 0.413 | 0.430 | 0 | 0 | 1.00 | 17.0 |
| AMR | haplotyp<br>e vs<br>pgs00354<br>2_hm_po<br>s_gr_ch3<br>8 | 1.00 | 0.417 | 0.409 | 0.426 | 0 | 0 | 1.00 | 17.0 |
| AMR | haplotyp<br>e vs<br>pgs00420<br>1_hm_po<br>s_gr_ch3<br>8 | 1.00 | 0.334 | 0.325 | 0.342 | 0 | 0 | 1.00 | 17.0 |
| AMR | haplotyp<br>e vs<br>pgs00420<br>2_hm_po<br>s_gr_ch3<br>8 | 1.00 | 0.319 | 0.310 | 0.328 | 0 | 0 | 1.00 | 17.0 |
| AMR | haplotyp<br>e vs<br>pgs00420<br>3_hm_po<br>s_gr_ch3<br>8 | 1.00 | 0.327 | 0.318 | 0.335 | 0 | 0 | 1.00 | 17.0 |
| AMR | haplotyp<br>e vs<br>pgs00420<br>4_hm_po<br>s_gr_ch3<br>8 | 1.00 | 0.331 | 0.322 | 0.340 | 0 | 0 | 1.00 | 17.0 |
| AMR | haplotyp<br>e vs<br>pgs00420<br>5_hm_po<br>s_gr_ch3<br>8 | 1.00 | 0.326 | 0.317 | 0.335 | 0 | 0 | 1.00 | 17.0 |
| CSA | haplotyp<br>e vs<br>pgs00031<br>3_hm_po<br>s_gr_ch3<br>8 | 0 | -0.0539 | -0.0608 | -0.0470 | 1.00 | 0 | 0 | 15.0 |
| CSA | haplotyp<br>e vs<br>pgs00066<br>7_hm_po<br>s_gr_ch3<br>8 | 0 | -0.0142 | -0.0210 | -0.00748 | 0.845 | 0.155 | 0 | 15.0 |
| CSA | haplotyp<br>e vs<br>pgs00068<br>9_hm_po<br>s_gr_ch3<br>8 | 0 | -0.181 | -0.188 | -0.174 | 1.00 | 0 | 0 | 15.0 |
| CSA | haplotyp<br>e vs<br>pgs00075<br>2_hm_po<br>s_gr_ch3<br>8 | 0.996 | 0.0109 | 0.00391 | 0.0177 | 0 | 0.421 | 0.579 | 15.0 |
| CSA | haplotyp<br>e vs<br>pgs00196<br>3_hm_po<br>s_gr_ch3<br>8 | 0.941 | 0.00635 | -<br>0.000315 | 0.0132 | 0 | 0.810 | 0.190 | 15.0 |
| CSA | haplotyp<br>e vs<br>pgs00218<br>1_hm_po<br>s_gr_ch3<br>8 | 0.447 | -<br>0.000568 | -0.00748 | 0.00629 | 0.0110 | 0.985 | 0.00425 | 15.0 |
| CSA | haplotyp<br>e vs<br>pgs00229<br>7_hm_po<br>s_gr_ch3<br>8 | 1.00 | 0.0521 | 0.0452 | 0.0589 | 0 | 0 | 1.00 | 15.0 |
| CSA | haplotyp<br>e vs<br>pgs00354<br>2_hm_po<br>s_gr_ch3<br>8 | 0 | -0.0373 | -0.0441 | -0.0305 | 1.00 | 0 | 0 | 15.0 |
| CSA | haplotyp<br>e vs<br>pgs00420<br>1_hm_po<br>s_gr_ch3<br>8 | 0 | -0.143 | -0.150 | -0.136 | 1.00 | 0 | 0 | 15.0 |
| CSA | haplotyp<br>e vs<br>pgs00420<br>2_hm_po<br>s_gr_ch3<br>8 | 0 | -0.141 | -0.148 | -0.134 | 1.00 | 0 | 0 | 15.0 |
| CSA | haplotyp<br>e vs<br>pgs00420<br>3_hm_po<br>s_gr_ch3<br>8 | 0 | -0.150 | -0.157 | -0.143 | 1.00 | 0 | 0 | 15.0 |
| CSA | haplotyp<br>e vs<br>pgs00420<br>4_hm_po<br>s_gr_ch3<br>8 | 0 | -0.146 | -0.153 | -0.139 | 1.00 | 0 | 0 | 15.0 |
| CSA | haplotyp<br>e vs<br>pgs00420<br>5_hm_po<br>s_gr_ch3<br>8 | 0 | -0.146 | -0.153 | -0.139 | 1.00 | 0 | 0 | 15.0 |
| EAS | haplotyp<br>e vs<br>pgs00031<br>3_hm_po<br>s_gr_ch3<br>8 | 0.707 | 0.00468 | -0.0102 | 0.0190 | 0.0528 | 0.677 | 0.270 | 8.00 |
| EAS | haplotyp<br>e vs<br>pgs00066<br>7_hm_po<br>s_gr_ch3<br>8 | 1.00 | 0.183 | 0.169 | 0.198 | 0 | 0 | 1.00 | 8.00 |
| EAS | haplotyp<br>e vs<br>pgs00068<br>9_hm_po<br>s_gr_ch3<br>8 | 0 | -0.0406 | -0.0552 | -0.0263 | 1.00 | 0.000500 | 0 | 8.00 |
| EAS | haplotyp<br>e vs<br>pgs00075<br>2_hm_po<br>s_gr_ch3<br>8 | 0 | -0.0727 | -0.0870 | -0.0578 | 1.00 | 0 | 0 | 8.00 |
| EAS | haplotyp<br>e vs<br>pgs00196<br>3_hm_po<br>s_gr_ch3<br>8 | 0.880 | 0.0106 | -0.00376 | 0.0250 | 0.00900 | 0.459 | 0.532 | 8.00 |
| EAS | haplotyp<br>e vs<br>pgs00218<br>1_hm_po<br>s_gr_ch3<br>8 | 0 | -0.116 | -0.130 | -0.101 | 1.00 | 0 | 0 | 8.00 |
| EAS | haplotyp<br>e vs<br>pgs00229<br>7_hm_po<br>s_gr_ch3<br>8 | 0.814 | 0.00776 | -0.00612 | 0.0221 | 0.0185 | 0.587 | 0.394 | 8.00 |
| EAS | haplotyp<br>e vs<br>pgs00354<br>2_hm_po<br>s_gr_ch3<br>8 | 0 | -0.135 | -0.149 | -0.120 | 1.00 | 0 | 0 | 8.00 |
| EAS | haplotyp<br>e vs<br>pgs00420<br>1_hm_po<br>s_gr_ch3<br>8 | 0 | -0.0555 | -0.0694 | -0.0411 | 1.00 | 0 | 0 | 8.00 |
| EAS | haplotyp<br>e vs<br>pgs00420<br>2_hm_po<br>s_gr_ch3<br>8 | 0 | -0.0516 | -0.0656 | -0.0373 | 1.00 | 0 | 0 | 8.00 |
| EAS | haplotyp<br>e vs<br>pgs00420<br>3_hm_po<br>s_gr_ch3<br>8 | 0 | -0.0476 | -0.0621 | -0.0333 | 1.00 | 0 | 0 | 8.00 |
| EAS | haplotyp<br>e vs<br>pgs00420<br>4_hm_po<br>s_gr_ch3<br>8 | 0 | -0.0392 | -0.0536 | -0.0251 | 1.00 | 0.000250 | 0 | 8.00 |
| EAS | haplotyp<br>e vs<br>pgs00420<br>5_hm_po<br>s_gr_ch3<br>8 | 0 | -0.0479 | -0.0621 | -0.0335 | 1.00 | 0 | 0 | 8.00 |
| EUR | haplotyp<br>e vs<br>pgs00031<br>3_hm_po<br>s_gr_ch3<br>8 | 1.00 | 0.124 | 0.122 | 0.126 | 0 | 0 | 1.00 | 557 |
| EUR | haplotyp<br>e vs<br>pgs00066<br>7_hm_po<br>s_gr_ch3<br>8 | 1.00 | 0.181 | 0.180 | 0.183 | 0 | 0 | 1.00 | 557 |
| EUR | haplotyp<br>e vs<br>pgs00068<br>9_hm_po<br>s_gr_ch3<br>8 | 1.00 | 0.116 | 0.114 | 0.117 | 0 | 0 | 1.00 | 557 |
| EUR | haplotyp<br>e vs<br>pgs00075<br>2_hm_po<br>s_gr_ch3<br>8 | 1.00 | 0.316 | 0.314 | 0.317 | 0 | 0 | 1.00 | 557 |
| EUR | haplotyp<br>e vs<br>pgs00196<br>3_hm_po<br>s_gr_ch3<br>8 | 1.00 | 0.288 | 0.287 | 0.290 | 0 | 0 | 1.00 | 557 |
| EUR | haplotyp<br>e vs<br>pgs00218<br>1_hm_po<br>s_gr_ch3<br>8 | 1.00 | 0.291 | 0.289 | 0.292 | 0 | 0 | 1.00 | 557 |
| EUR | haplotyp<br>e vs<br>pgs00229<br>7_hm_po<br>s_gr_ch3<br>8 | 1.00 | 0.173 | 0.172 | 0.175 | 0 | 0 | 1.00 | 557 |
| EUR | haplotyp<br>e vs<br>pgs00354<br>2_hm_po<br>s_gr_ch3<br>8 | 1.00 | 0.281 | 0.279 | 0.282 | 0 | 0 | 1.00 | 557 |
| EUR | haplotyp<br>e vs<br>pgs00420<br>1_hm_po<br>s_gr_ch3<br>8 | 1.00 | 0.0783 | 0.0768 | 0.0798 | 0 | 0 | 1.00 | 557 |
| EUR | haplotyp<br>e vs<br>pgs00420<br>2_hm_po<br>s_gr_ch3<br>8 | 1.00 | 0.0793 | 0.0777 | 0.0808 | 0 | 0 | 1.00 | 557 |
| EUR | haplotyp<br>e vs<br>pgs00420<br>3_hm_po<br>s_gr_ch3<br>8 | 1.00 | 0.0771 | 0.0756 | 0.0786 | 0 | 0 | 1.00 | 557 |
| EUR | haplotyp<br>e vs<br>pgs00420<br>4_hm_po<br>s_gr_ch3<br>8 | 1.00 | 0.0778 | 0.0764 | 0.0793 | 0 | 0 | 1.00 | 557 |
| EUR | haplotyp<br>e vs<br>pgs00420<br>5_hm_po<br>s_gr_ch3<br>8 | 1.00 | 0.0791 | 0.0776 | 0.0806 | 0 | 0 | 1.00 | 557 |

**eTable 5:** Performance of the LPA-haplotype model in predicting Lp(a) concentrations. Values are posterior medians with 95% credible intervals.

| ancestry | metric | threshold | estimate | lower | upper |
| --- | --- | --- | --- | --- | --- |
| Overall | accuracy | 125 | 0.796 | 0.600 | 0.894 |
| EUR | accuracy | 125 | 0.869 | 0.783 | 0.909 |
| AMR | accuracy | 125 | 0.807 | 0.678 | 0.886 |
| AFR | accuracy | 125 | 0.722 | 0.609 | 0.816 |
| Overall | f_meas | 125 | 0.624 | 0.434 | 0.760 |
| EUR | f_meas | 125 | 0.692 | 0.580 | 0.757 |
| AMR | f_meas | 125 | 0.575 | 0.389 | 0.713 |
| AFR | f_meas | 125 | 0.604 | 0.499 | 0.696 |
| Overall | lr_neg | 125 | 0.523 | 0.338 | 0.730 |
| EUR | lr_neg | 125 | 0.423 | 0.337 | 0.552 |
| AMR | lr_neg | 125 | 0.566 | 0.394 | 0.765 |
| AFR | lr_neg | 125 | 0.571 | 0.446 | 0.691 |
| Overall | lr_pos | 125 | 8.89 | 2.95 | 17.4 |
| EUR | lr_pos | 125 | 12.7 | 5.56 | 21.4 |
| AMR | lr_pos | 125 | 7.91 | 2.75 | 16.6 |
| AFR | lr_pos | 125 | 8.15 | 3.35 | 14.8 |
| Overall | npv | 125 | 0.788 | 0.559 | 0.903 |
| EUR | npv | 125 | 0.877 | 0.765 | 0.922 |
| AMR | npv | 125 | 0.821 | 0.658 | 0.906 |
| AFR | npv | 125 | 0.670 | 0.527 | 0.804 |
| Overall | ppv | 125 | 0.807 | 0.596 | 0.895 |
| EUR | ppv | 125 | 0.822 | 0.687 | 0.891 |
| AMR | ppv | 125 | 0.777 | 0.513 | 0.885 |
| AFR | ppv | 125 | 0.839 | 0.689 | 0.909 |
| Overall | sens | 125 | 0.511 | 0.331 | 0.682 |
| EUR | sens | 125 | 0.598 | 0.478 | 0.679 |
| AMR | sens | 125 | 0.470 | 0.296 | 0.630 |
| AFR | sens | 125 | 0.465 | 0.363 | 0.579 |
| Overall | spec | 125 | 0.944 | 0.845 | 0.968 |
| EUR | spec | 125 | 0.953 | 0.895 | 0.972 |
| AMR | spec | 125 | 0.942 | 0.844 | 0.970 |
| AFR | spec | 125 | 0.943 | 0.867 | 0.967 |
| Overall | accuracy | 150 | 0.814 | 0.643 | 0.899 |
| EUR | accuracy | 150 | 0.862 | 0.766 | 0.906 |
| AMR | accuracy | 150 | 0.836 | 0.709 | 0.909 |
| AFR | accuracy | 150 | 0.754 | 0.647 | 0.855 |
| Overall | f_meas | 150 | 0.557 | 0.409 | 0.685 |
| EUR | f_meas | 150 | 0.574 | 0.466 | 0.675 |
| AMR | f_meas | 150 | 0.545 | 0.366 | 0.674 |
| AFR | f_meas | 150 | 0.555 | 0.448 | 0.663 |
| Overall | lr_neg | 150 | 0.599 | 0.453 | 0.752 |
| EUR | lr_neg | 150 | 0.571 | 0.460 | 0.683 |
| AMR | lr_neg | 150 | 0.612 | 0.467 | 0.786 |
| AFR | lr_neg | 150 | 0.613 | 0.493 | 0.726 |
| Overall | lr_pos | 150 | 11.1 | 3.26 | 22.8 |
| EUR | lr_pos | 150 | 13.7 | 4.63 | 25.8 |
| AMR | lr_pos | 150 | 11.2 | 3.32 | 26.4 |
| AFR | lr_pos | 150 | 10.8 | 3.54 | 21.3 |
| Overall | npv | 150 | 0.802 | 0.598 | 0.904 |
| EUR | npv | 150 | 0.870 | 0.756 | 0.918 |
| AMR | npv | 150 | 0.839 | 0.684 | 0.916 |
| AFR | npv | 150 | 0.717 | 0.576 | 0.838 |
| Overall | ppv | 150 | 0.804 | 0.594 | 0.904 |
| EUR | ppv | 150 | 0.806 | 0.642 | 0.900 |
| AMR | ppv | 150 | 0.797 | 0.557 | 0.909 |
| AFR | ppv | 150 | 0.822 | 0.652 | 0.917 |
| Overall | sens | 150 | 0.426 | 0.296 | 0.565 |
| EUR | sens | 150 | 0.450 | 0.351 | 0.555 |
| AMR | sens | 150 | 0.414 | 0.259 | 0.551 |
| AFR | sens | 150 | 0.413 | 0.318 | 0.527 |
| Overall | spec | 150 | 0.962 | 0.876 | 0.980 |
| EUR | spec | 150 | 0.967 | 0.906 | 0.982 |
| AMR | spec | 150 | 0.964 | 0.886 | 0.984 |
| AFR | spec | 150 | 0.962 | 0.886 | 0.980 |
| Overall | accuracy | 175 | 0.817 | 0.531 | 0.917 |
| EUR | accuracy | 175 | 0.880 | 0.732 | 0.924 |
| AFR | accuracy | 175 | 0.774 | 0.620 | 0.890 |
| Overall | f_meas | 175 | 0.505 | 0.319 | 0.695 |
| EUR | f_meas | 175 | 0.519 | 0.374 | 0.664 |
| AFR | f_meas | 175 | 0.490 | 0.339 | 0.653 |
| Overall | lr_neg | 175 | 0.662 | 0.443 | 1.31 |
| EUR | lr_neg | 175 | 0.632 | 0.475 | 0.773 |
| AFR | lr_neg | 175 | 0.673 | 0.493 | 0.811 |
| Overall | lr_pos | 175 | 11.9 | 0.729 | 31.0 |
| EUR | lr_pos | 175 | 19.3 | 5.40 | 39.2 |
| AFR | lr_pos | 175 | 12.0 | 3.52 | 27.7 |
| Overall | npv | 175 | 0.812 | 0.521 | 0.919 |
| EUR | npv | 175 | 0.884 | 0.730 | 0.929 |
| AFR | npv | 175 | 0.762 | 0.591 | 0.889 |
| Overall | ppv | 175 | 0.810 | 0.578 | 0.909 |
| EUR | ppv | 175 | 0.815 | 0.651 | 0.900 |
| AFR | ppv | 175 | 0.818 | 0.637 | 0.909 |
| Overall | sens | 175 | 0.369 | 0.211 | 0.574 |
| EUR | sens | 175 | 0.383 | 0.252 | 0.536 |
| AFR | sens | 175 | 0.350 | 0.226 | 0.523 |
| Overall | spec | 175 | 0.970 | 0.460 | 0.987 |
| EUR | spec | 175 | 0.981 | 0.930 | 0.989 |
| AFR | spec | 175 | 0.971 | 0.908 | 0.986 |
| Overall | accuracy | 200 | 0.837 | 0.546 | 0.933 |
| EUR | accuracy | 200 | 0.900 | 0.773 | 0.939 |
| AFR | accuracy | 200 | 0.795 | 0.615 | 0.906 |
| Overall | f_meas | 200 | 0.460 | 0.263 | 0.679 |
| EUR | f_meas | 200 | 0.493 | 0.329 | 0.656 |
| AFR | f_meas | 200 | 0.420 | 0.268 | 0.621 |
| Overall | lr_neg | 200 | 0.698 | 0.443 | 0.944 |
| EUR | lr_neg | 200 | 0.656 | 0.491 | 0.803 |
| AFR | lr_neg | 200 | 0.732 | 0.526 | 0.877 |
| Overall | lr_pos | 200 | 11.2 | 1.25 | 35.1 |
| EUR | lr_pos | 200 | 23.6 | 4.41 | 51.2 |
| AFR | lr_pos | 200 | 10.5 | 2.39 | 27.6 |
| Overall | npv | 200 | 0.841 | 0.556 | 0.935 |
| EUR | npv | 200 | 0.905 | 0.765 | 0.943 |
| AFR | npv | 200 | 0.798 | 0.636 | 0.906 |
| Overall | ppv | 200 | 0.780 | 0.536 | 0.897 |
| EUR | ppv | 200 | 0.804 | 0.614 | 0.899 |
| AFR | ppv | 200 | 0.772 | 0.547 | 0.890 |
| Overall | sens | 200 | 0.332 | 0.166 | 0.579 |
| EUR | sens | 200 | 0.356 | 0.223 | 0.519 |
| AFR | sens | 200 | 0.292 | 0.175 | 0.491 |
| Overall | spec | 200 | 0.971 | 0.769 | 0.989 |
| EUR | spec | 200 | 0.985 | 0.926 | 0.992 |
| AFR | spec | 200 | 0.972 | 0.886 | 0.988 |
| Overall | rsq | NA | 0.459 | 0.315 | 0.596 |
| EUR | rsq | NA | 0.530 | 0.382 | 0.652 |
| EAS | rsq | NA | 0.402 | 0.201 | 0.571 |
| CSA | rsq | NA | 0.437 | 0.258 | 0.593 |
| AMR | rsq | NA | 0.471 | 0.334 | 0.616 |
| AFR | rsq | NA | 0.450 | 0.334 | 0.587 |

**eTable 6:** Number needed to test to identify one individual with elevated Lp(a). Values are posterior medians with 95% credible intervals.

| ancestry | threshold | nnt | lower | upper |
| --- | --- | --- | --- | --- |
| Overall | 125 | 1.24 | 1.12 | 1.68 |
| EUR | 125 | 1.22 | 1.12 | 1.46 |
| AMR | 125 | 1.29 | 1.13 | 1.95 |
| AFR | 125 | 1.19 | 1.10 | 1.45 |
| Overall | 150 | 1.24 | 1.11 | 1.68 |
| EUR | 150 | 1.24 | 1.11 | 1.56 |
| AMR | 150 | 1.25 | 1.10 | 1.80 |
| AFR | 150 | 1.22 | 1.09 | 1.53 |
| Overall | 175 | 1.23 | 1.10 | 1.73 |
| EUR | 175 | 1.23 | 1.11 | 1.54 |
| AFR | 175 | 1.22 | 1.10 | 1.57 |
| Overall | 200 | 1.28 | 1.11 | 1.87 |
| EUR | 200 | 1.24 | 1.11 | 1.63 |
| AFR | 200 | 1.30 | 1.12 | 1.83 |

