## Supplement for "Genetic Prediction of Circulating Lipoprotein(a) Levels in Diverse Populations"

### Supplemental Methods: LPA Haplotype Model Development

#### Overview

We developed a software tool that enables prediction of Lp(a) levels using only genotypes of common and low-frequency SNPs at the LPA locus. This approach maps SNP haplotypes, representing LPA alleles, to Lp(a) levels predicted by our previously developed sequencing-based model. Because SNP haplotype information can be readily generated from many forms of genetic data (including SNP-array data and low-coverage sequencing data), this approach can be applied to most existing genetic data sets, circumventing the need to genotype complex structural variation at LPA. A similar approach has been successfully applied within the Icelandic population.^1^

For SNP haplotype-based prediction to work well, haplotypes of target individuals must be represented among the reference haplotypes used to train the model, and the SNP haplotypes must be specific enough to tag rare LPA alleles. The All of Us (AoU) data set was ideal for building this predictor given its diversity and scale.

#### LPA SNP Selection

To identify SNPs at the LPA locus expected to be broadly available and accurately genotyped in data sets derived from either whole-genome sequencing or imputation, we started with biallelic SNPs observed in the 1000 Genomes Phase 3 data set (1000 Genomes Consortium 2015), lifted to GRCh38, and restricted to SNPs also present in the HRC v1.1 reference panel,^2^ TOPMed-r2 reference panel,^3^ UK Biobank N=200K WGS imputation panel,^4^ AoU ACAF variant call set,^5^ and gnomAD v4.1 data set.^6^

We excluded SNPs annotated in gnomAD as failing QC (FILTER ≠ PASS) or falling in low-complexity regions or segmental duplications, and SNPs falling within structural variants in the gnomAD v4.1 SV call set with FILTER=PASS and allele frequency >0.001. We further restricted to SNPs with AAscore >0.9 and FILTER=PASS in the UKB N=200K WGS data set, call rate >0.99 at GQ ≥ 20 in the AoU WGS data set, and high imputation accuracy (R² >0.9 in TOPMed-imputed data sets and INFO >0.8 in the UKB imp_v3 data set). Finally, we required that allele frequencies in UKB WGS-derived and imputed data sets closely match (UKB N=200K WGS AF and TOPMed-imputed AF within 0.002; imp_v3 AF and TOPMed-imputed AF within 0.001).

These filters resulted in a set of 1,408 common and low-frequency SNPs at the LPA locus (chr6:160.4–160.8 Mb in GRCh38, containing the LPA gene and ~130 kb on each flank).

#### LPA SNP Haplotypes in All of Us

To generate phased SNP haplotypes of AoU participants, we extracted the selected SNPs from the ACAF variant call set using bcftools v1.21^7^ to split and normalize multi-allelic variants and set low-quality genotypes (GQ <20) to missing. We then phased these SNPs onto the SNP-array scaffold using SHAPEIT v5.1.1 phase_common.^4^ We converted phased SNP haplotypes to plain-text format using plink2 –recode phylip-phased and merged the SNP haplotype data with estimates of Lp(a) produced by each haplotype, generated by applying an updated version of our sequencing-based Lp(a) prediction model to AoU whole-genome sequencing data.^8,9^

#### SNP-Haplotype Model for Lp(a)

To build a mapping from SNP haplotypes to estimated Lp(a) levels, we encoded SNP haplotypes into a bifurcating tree, similar in spirit to Fig. 4 of Boettger et al. 2016.^10^ We first ordered the selected SNPs by increasing distance from the KIV-2 repeat, prioritizing SNPs within the LPA gene before flanking SNPs. We then added each haplotype in turn to a binary tree as follows: starting at the root node, for each SNP in turn (in the defined order), we moved from the current node to its child corresponding to the haplotype’s allele at that SNP. At each node, we maintained running totals of (i) the number of haplotypes passing through the node, (ii) the sum of Lp(a) estimates for those haplotypes, and (iii) the sum of squares of the Lp(a) estimates. We ensured that each individual was counted only once per node (i.e., until an individual’s two haplotypes diverged, we updated totals for each visited node only once).

To output the model, we performed a depth-first traversal through the SNP haplotype tree, visiting only nodes corresponding to (sub-)haplotypes present in >20 AoU participants. For each such node, we recorded the mean and standard deviation of the Lp(a) estimates of contributing haplotypes. We rounded mean Lp(a) estimates in nmol/L to the nearest integer and standard deviations to the nearest multiple of 5. We restricted to haplotypes with >20 carriers to ensure compliance with the AoU Data and Statistics Dissemination policy for public release. No phenotype information for AoU participants was used to train the model; we only used sequencing data to estimate the amounts of Lp(a) produced by LPA alleles observed in AoU.

To predict Lp(a) produced by a target haplotype, our tool proceeds down the stored bifurcating tree according to the allele observed at each SNP in turn until the SNP haplotype path can no longer be extended (because doing so would reach a node observed in ≤20 AoU participants). The mean and standard deviation of Lp(a) recorded for the final visited node are then reported as the prediction for the target haplotype.

### Penn Medicine BioBank Banner Author List and Contribution Statements

#### PMBB Leadership Team

Daniel J. Rader, M.D., Marylyn D. Ritchie, Ph.D.

**Contribution**: All authors contributed to securing funding, study design and oversight. All authors reviewed the final version of the manuscript.

#### Patient Recruitment and Regulatory Oversight

JoEllen Weaver, Nawar Naseer, Ph.D., M.P.H., Giorgio Sirugo, M.D., P.h.D., Afiya Poindexter, Jenna Dever, Aidan Harvey, Sydney Linn, Naman Srivastava

**Contributions**: JW manages patient recruitment and regulatory oversight of study. NN manages participant engagement, assists with regulatory oversight, and researcher access. GS assists with researcher access. AP, JD, AH, SL, and NS perform recruitment and enrollment of study participants.

#### Lab Operations

JoEllen Weaver, Meghan Livingstone, Fred Vadivieso, Stephanie DerOhannessian, Teo Tran, Julia Stephanowski, Salma Santos, Ned Haubein, P.h.D., Joseph Dunn

**Contribution**: JW, ML, FV, SD conduct oversight of lab operations. ML, FV, AK, SD, TT, JS, SS perform sample processing. NH, JD are responsible for sample tracking and the laboratory information management system.

#### Clinical Informatics

Anurag Verma, Ph.D., Colleen Morse Kripke, M.S. DPT, MSA, Marjorie Risman, M.S., Renae Judy, B.S., Colin Wollack, M.S.

**Contribution**: All authors contributed to the development and validation of clinical phenotypes used to identify study subjects and (when applicable) controls.

#### Genome Informatics

Anurag Verma Ph.D., Shefali S. Verma, Ph.D., Scott Damrauer, M.D., Yuki Bradford, M.S., Scott Dudek, M.S., Theodore Drivas, M.D., Ph.D., Zachary Rodriguez, Ph.D.

**Contribution**: AV, SSV, and SD are responsible for the analysis, design, and infrastructure needed to quality control genotype and exome data. YB performs the analysis. TD and AV provides variant and gene annotations and their functional interpretation of variants.

For PMBB, please use:
For Regeneron, please use:
